# A Predictive Modelling Framework for COVID-19 Transmission to Inform the Management of Mass Events

**DOI:** 10.1101/2021.05.13.21256857

**Authors:** Claire Donnat, Freddy Bunbury, Jack Kreindler, Filippos T. Filippidis, Austen El-Osta, Tõnu Esko, Matthew Harris

**Affiliations:** Department of Statistics, University of Chicago, Chicago, USA; Department of Plant Biology, Carnegie Institution for Science, Stanford, USA; School of Public Health, Imperial College, London, UK; Institute of Genomics, University of Tartu, Tartu, Estonia

## Abstract

Modelling COVID-19 transmission at live events and public gatherings is essential to evaluate and control the probability of subsequent outbreaks. Model estimates can be used to inform event organizers about the possibility of super-spreading and the predicted efficacy of safety protocols, as well as to communicate to participants their personalised risk so that they may choose whether to attend. Yet, despite the fast-growing body of literature on COVID transmission dynamics, current risk models either neglect contextual information on vaccination rates or disease prevalence or do not attempt to quantitatively model transmission, thus limiting their potential to provide insightful estimates. This paper attempts to bridge this gap by providing informative risk metrics for live public events, along with a measure of their associated uncertainty. Starting with a thorough review of the literature and building upon existing models, our approach ties together three main components: (a) reliable modelling of the number of infectious cases at the time of the event, (b) evaluation of the efficiency of pre-event screening and risk mitigation protocols, and (c) modelling the transmission dynamics during the event. We demonstrate how uncertainty in the input parameters can be included in the model using Monte Carlo simulations. We discuss the underlying assumptions and limitations of our approach and implications for policy around live events management.

## 1. Introduction

More than a year after a global, unprecedented cancellation of live events in March 2020, the future of live events and the entertainment industry remains uncertain despite increasing vaccination rates and low community prevalence levels (at the time of writing). The main concern raised by these gatherings lies in their susceptibility to “super-spreading”—a scenario whereby a few contagious participants inadvertently infect a disproportionately large number of others [1, 2, 3, 4, 5, 6]. The role of super-spreading events has been highlighted as a significant driver of the pandemic [7, 8, 9, 10, 11]. Despite the planned re-opening of live events in the UK on June 21^st^ 2021, the threat of existing and emergent COVID-19 variants coupled Preprint. Under review. to dwindling immunity from vaccination over time suggests that policy makers and event organizers will likely continue to struggle with the following two questions: (a) Is the COVID-19 transmission risk posed by these events tolerable? and (b) What additional safety measures can be feasibly deployed to reduce this risk? The answer to these questions is inherently tied to the estimation of two quantities: *the number of infections occurring at the event*, and *the post-event secondary attack rate*, or number of subsequent infections in the participants’ social circles. Evaluating the safety (or lack thereof) of large public gatherings can then be reframed as quantifying the significance and magnitude of their effect on the distribution of the number of primary and secondary COVID-19 cases. Yet, despite the growing body of literature on COVID-19 risk evaluation and recent efforts to evaluate the safety of live events, their effect on COVID-19 transmission remains ill-characterized. Nevertheless, over the past several months, a number of calculators were developed to estimate this risk [12, 13, 14, 15]. These methods can typically be placed in one of three categories.

a. *Ranking heuristics:* These estimators typically rank events on a scale ranging from “low” to “high” risk based on the feedback of medical experts [13, 16, 17, 18]. However, these heuristics do not take into account any contextual information, including the prevalence, such that the risk associated with the event would be classified as high regardless of whether it was held in August 2020 (background prevalence of 1 in 3,000 individuals) or January 2021 (prevalence of 1 in 60 individuals^1^).
b. *Context-based heuristics:* These calculators estimate the probability of encountering one COVID-case based on the number of people attending an event [12, 13]. Whilst the latter are more context-aware than risk assessment charts, they do not account for modelling transmission dynamics—which is undeniably one of the main unknowns in the spread of viral epidemics — and consequently rarely stratify risk by type of activity. To exemplify, a classical music recital of 1.5 hours for the BBC Proms would potentially be considered equally risky to a 3-hour concert in which participants could be expected to sing along.
c. *Transmission risk calculators:* Stemming from the physics of fluid dynamics, these calculators focus on modelling the aerosolization and spread of micro-droplets — typically in a closed/indoor environment [19, 20, 21, 22]. These fine-grained models have to be combined with extensive simulations of crowd movements in order to model transmission dynamics during any given event.

Regardless of their category, most of these models rely on a large number and wide range of input parameters, including (but not restricted to) the prevalence of the disease. Whilst certain calculators attempt to bridge the gap between expert heuristics and physical models [12, 23], they are not capable of predicting the risk of a future event. Moreover, all of these estimators provide point estimates − in other words, their output is a single number to quantify the risk. Given the uncertainty associated with all the inputs and the parametrization of the problem, the provision of a single consolidated outcome or number can potentially be misleading. This is because a singular focus on the expected outcome precludes consideration of the distribution of all possible outcomes, including worst-case scenarios.

### Mitigating transmission risk

Meanwhile, with the increasing vaccination rates in several countries around the world, a few initiatives have begun to evaluate the outbreak risk associated with live events [24, 25, 26, 27]. This is because vaccinated individuals may still be infected with SARS-CoV-2 [28, 29], and even antigen-test based screening of ticket holders offers no guarantee due to false negatives [30, 31]. The estimation of what constitutes an admissible level of risk thus poses a difficult conundrum to the live event industry. To begin answering these questions, the CAPACITY study [32] — a partnership between CERTIFIC (a private, remote testing, health status and identify certification service), LiveNation (a live events production company) and Imperial College London − aims to predict and measure the outcomes of full capacity live events whilst ensuring rigorous implementation and alignment to current public health and recommended safety measures. Central to this study is the provision of a streamlined and efficient pre-event screening protocol of all ticket holders using professionally-witnessed rapid at-home antigen tests followed by post-event monitoring based on antigen tests, surveys and safety recommendations (see Appendix D). In this setting, providing risk estimates not only becomes essential in communicating to the ticket holders their own level of risk so that they may make an informed decision of whether or not to attend the event, but also necessary to inform event managers and policy makers on the likelihood of an outbreak − a task which serves here as the motivating application behind this paper.

### A working example: Concert at the Royal Albert Hall

In order to understand and illustrate the potential challenges that arise in the risk estimation for the CAPACITY study, we consider as an example a concert at the Royal Albert Hall (RAH), and demonstrate how to estimate the associated risk assuming a near-capacity attendance of 5,000 in the main concert hall, which has a volume of 86,650 m^3^ [33], with a dwell time of 3 hours. Attendees will be assumed to be a cross-section representative of the general British public and will be required to have a negative COVID antigen test result within 2 days prior to the event, as well as satisfying other self-declared symptoms and exposure-risk questions. Vaccination status would be requested, but not required for attendance, and full compliance with mask wearing is assumed in our default example.

### Goals and Contributions

The objectives of our modelling approach are three-fold: (a) enable the quantitative comparison of different activities and event characteristics, (b) estimate the efficacy of various safety protocols and (c) provide a predictive risk assessment, i.e, the risk associated with a scheduled future event. To this end, we delineate our approach into three sequential steps (see Fig. 1) as follows: 1) estimating the number of contagious participants, 2) evaluating the transmission dynamics, and 3) comparing the risk of holding the event with the null model i.e. if the event had not taken place. We illustrate the application of our risk modelling pipeline in the RAH example to highlight the risk’s dependency on factors such as prevalence, mask wearing, number of attendees and event duration. In particular, we demonstrate how this particular event held on three different dates (3^rd^ August 2020, 18^th^ January 2021, 8^th^ March 2021) would likely lead to transmission events only slightly above background rates (0.5 vs 0.2, 6.7 vs 3.5, and 5.4 vs 2.5, respectively (Table 2)). However, the 97.5 percentile of the prediction interval for the infections at the event would likely be substantially higher than the background rate (6.8 vs 2, 89 vs 8, and 71 vs 7, respectively), further demonstrating that sole reliance on vaccination and antigen testing to gain entry would significantly underestimate the tail risk of the event. However, we emphasize that the goal of this paper is not to present a novel “state-of-the-art” risk estimation procedure because COVID-19 transmission mechanisms remain poorly characterized, and we acknowledge that our approach requires certain simplifications and assumptions which we discuss at length in the last section of this paper. Rather, faced with the need to provide a risk evaluation tool despite many unknowns, our estimation pipeline combines the best tools at hand to assess the order of magnitude of the risk − thereby opening the avenue for further work on contextualized COVID risk assessment. Our model can be applied to any event occurring in the near future, and is presented in a user-friendly R Shiny interface^2^.

**Figure 1:**
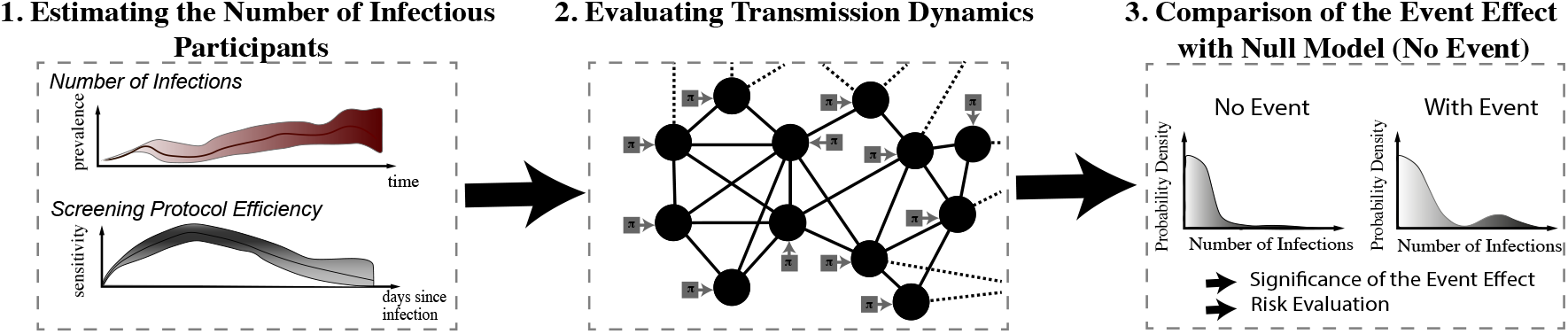
Summary of our modelling pipeline

## 2. Modelling The Risk of a Large Public Event

### Step 1. Estimating the Number of Infectious Participants

#### Projected Incidence

The first step in our risk modelling procedure is to predict the number of infectious cases attending a given future event. COVID forecasting is undeniably an involved task, as reflected by its impressive corresponding body of literature (e.g, agent-based models, or **S**usceptible-**E**xposed-**I**nfectious-**R**emoved (SEIR) models [34, 35, 36, 37, 38, 39, 40, 41, 42, 43, 44]). Predicting the number of new cases per day typically depends on the choice of a specific parameterization (e.g, an exponential growth for computing the reproductive number *R* [45, 46], etc), whose validity is severely hindered by continuous updates in public policies. To alleviate these concerns, we use a non-parametric k-nearest neighbour (kNN) approach. Using all trajectories of the disease incidence across countries and time since the beginning of the pandemic, we compute the k = 100 closest trajectories (in terms of the 𝓁_2_ loss) on time windows of four weeks. These nearest neighbours’ subsequent trajectories are then used to predict the daily incidence rate in the days leading to the event. We defer to Appendix A for a more in-depth discussion of this estimation procedure, as well as an evaluation of its performance compared to more standard methods. Figure 2 presents a comparison of the projected incidence for various randomly sampled dates (August 3^rd^ 2020, January 18^th^ 2021, and March 8^th^ 2021) for the RAH concert using four weeks of fitting, and predicting four weeks in advance. Note in particular the good coverage provided by our method (the convex hull of the 95% prediction intervals for the projected incidences contains the actual observations). These plots highlight the importance and variability of the incidence, which varied by orders of magnitude between August 2020 and January 2021.

**Figure 2:**
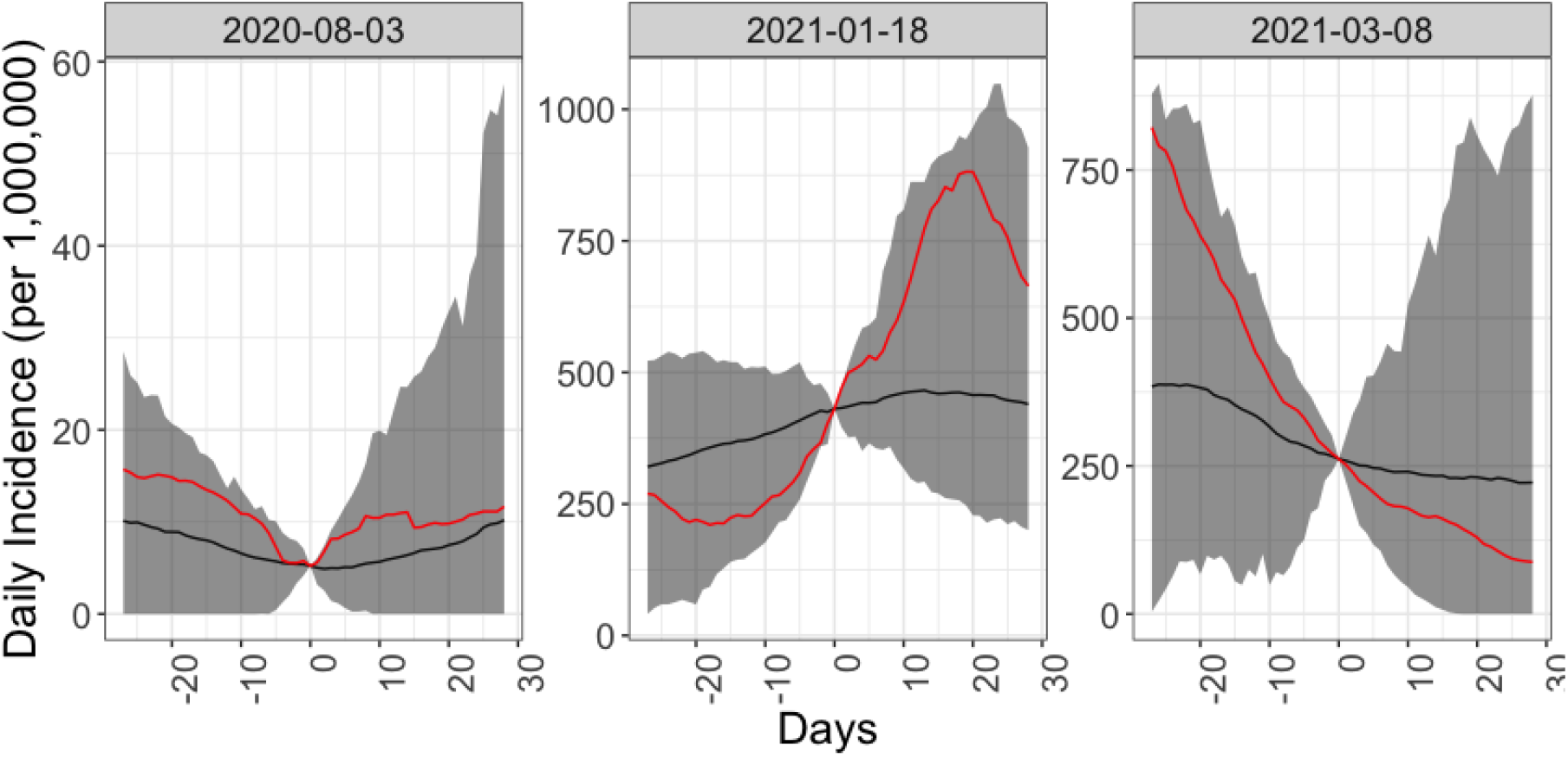
Projected incidence (average and 95% prediction interval). The 4 weeks of fitting is denoted in light gray, and the confidence interval for the next for weeks is highlighted in dark gray. We note that our kNN method provides good coverage (the observed trajectory lies well within the 95% prediction interval).

#### Under-ascertainment Bias

The estimated number of new cases based on official incidence data will then need to be corrected for under-ascertainment. The latter refers to the downward bias of the reported prevalence in the population, due for instance to limited testing capacity, low test sensitivity or people being unwilling or unable to take a test. To this end, we compare the ratio of the number of deaths over reported cases (translated by three weeks) to an expected, age-stratified Infection-Fatality Ratio^3^ (see Appendix A for more details). To highlight the potential importance of this correction step, the ascertainment rate for the United Kingdom was evaluated as over 90% for August 2020, but below 40% for December 2020.

#### Determining the Number of Infectious participants at the event

Having predicted the background daily incidence rate, we turn to the estimation of the number of infectious participants who will attend the event despite the screening protocols. For an infectious individual to attend the event in spite of the CAPACITY study’ screening protocol, they must (a) have no COVID-like symptoms or fail to report them on the morning of the event, (b) receive a (false) negative result during antigen testing D = 2 days prior to the event, and (c) be contagious (rather than simply infected) at the time of the event. We evaluate the joint probability of these events as follows, and, for the sake of clarity, refer the reader to Appendix A for an in-depth explanation of our estimation procedure.

##### (a) Symptoms-Check Failure

One of the main challenges associated with the COVID-19 crisis is the number of asymptomatic cases - that is, infected individuals that do not express symptoms and are thus unaware of their potential infectiousness. This group includes individuals that are either pre-symptomatic or completely asymptomatic during the course of their illness − the latter are estimated to represent roughly 25% of all cases [47]. For symptomatic patients, the probability of having symptoms on the day of the event is also a function of time since infection. To account for this temporal dependency, we use estimates of the incubation period (defined as the number of days between infection and symptom onset) from McAloon et al. [48] and data on symptoms duration from van Kampen et al. [49] to estimate the probability for a ticket holder infected *k* days before the event to exhibit symptoms on the day of the event. A density plot of this probability is displayed in red in Figure 3a.

**Figure 3:**
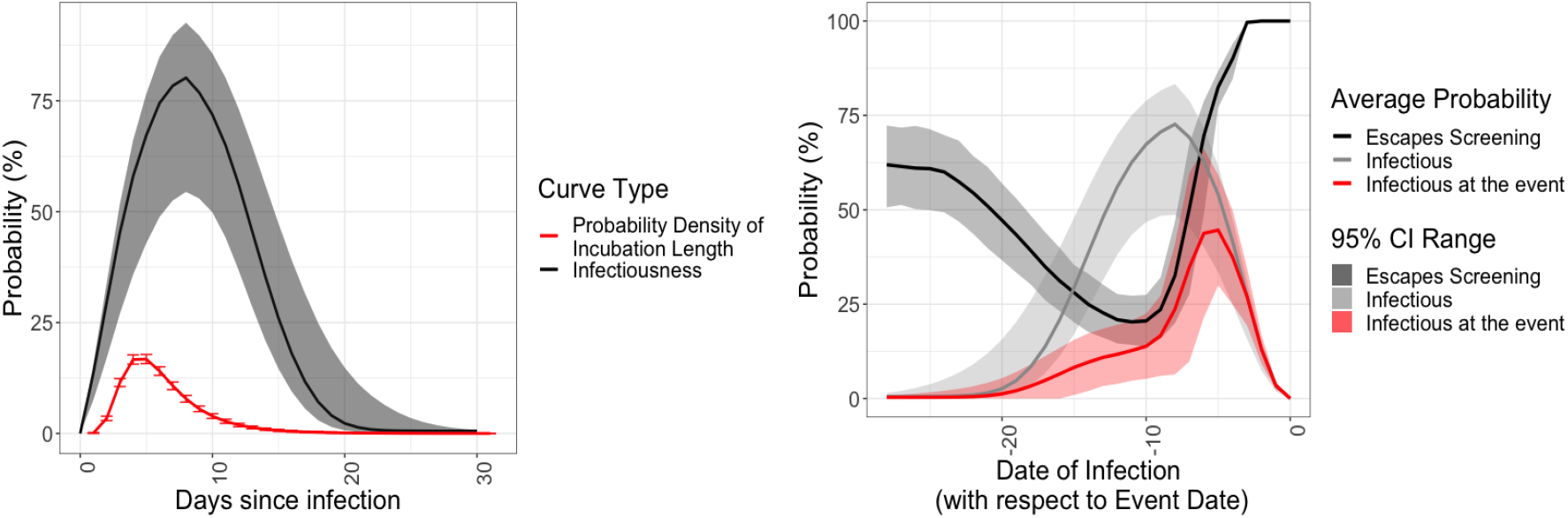
(a) Density of the COVID-19 incubation time, and Percentage culture positive. (b) Probability that an individual is infectious (light grey),that the screening protocol will miss them (black), and that they will be missed and so attend the event (red), as a function of days since infection. The shaded regions denote the uncertainty of this estimate due to the uncertainty on the sensitivity of the test. The distribution of the incubation time already integrates the uncertainty on the parameters *μ* and *σ* of the log-normal distribution.

##### (b) Antigen test failure

The sensitivity of COVID tests depends heavily on the time since infection — whether these are the gold-standard PCR or Lateral Flow Antigen Assays [50]. Moreover, studies have shown that LFA tests have much lower sensitivity on asymptomatic individuals than symptomatic: in particular, according to a recent CDC report [51], Rapid Antigen testing has 80% sensitivity on symptomatic individuals, but only 40% sensitivity on asymptomatic individuals. Coupling the sensitivity estimates [50, 51] with the distribution of incubation period and estimated percentage of asymptomatic cases [48, 47], for each individual infected at day *k* taking an antigen test *D* days before the event, the probability of getting through the filtering protocol is thus given by the formula:

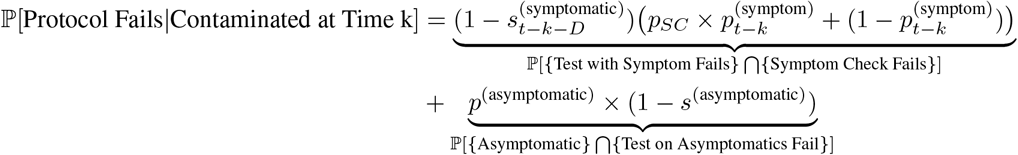

where 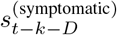 and *s*^(asymptomatic)^ are respectively the sensitivities of the test taken *D* days before the event for a symptomatic participant infected *t* − *k* days before the event and an asymptomatic individual. The parameter 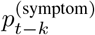 denotes the probability for a symptomatic individual to exhibit symptoms *t* − *k* days after infection, hereas *p*^(asymptomatic)^ infection, is the probability of being asymptomatic. Finally, the variable denotes the probability of the Symptoms Check failing — namely, that the participant does not want to report their symptoms (see Appendix A for more details). The curve in black on Figure 3b shows the probability of the failure of the screening protocol as a function of days after infection. The shaded areas denote the uncertainty around this estimate due to the variability of the incubation time.

##### (c) Infectiousness

The infectiousness of the participants — that is, the propensity of an infected ticket holder to contaminate others — is a function of time since infection. In order to estimate this relationship, we build upon the existing literature studying the link between RT-PCR thresholds and cultivable virus [52, 53]. The percentage of culturable viral material in the sample can indeed be used as a proxy for infectiousness. Using the estimated percentages of viable samples [52, 53] as a function of time since symptom onset, compounded with distribution of the incubation period duration [48], we compute an estimate of the infectiousness as a function of time since infection (black curve in Figure 3a). A more complete description of this estimation procedure is presented in Appendix A. The results are presented in Figure 3b. The red line in Figure 3b shows the resulting probability for an infectious ticket holder to pass through the screening protocol and be allowed into the event. Note that ticket holders that have been infected five days before the event are the most likely to be infectious and let in the venue on the day of the event.

#### Determining the number of participants at risk

Finally, the last quantity that we need to infer before getting into the specifics of the transmission mechanisms is the number of participants at risk of being infected who present at the event. This requires a knowledge of the participants’ COVID susceptibility status, i.e, has the participant already had COVID in the previous year and/or has the participant been vaccinated? While previous history could be imputed through additional questions (e.g., previous positive test for COVID, symptoms, etc, combined in a model such as in [54]), for the sake of simplicity, we only consider the vaccination status of the participants — thus leaving out the proportion of the population that had COVID but was not yet vaccinated. This induces a risk estimate that is biased upward, and is thus more conservative. We impute missing data (cases where the participants have not filled in their vaccination status) using linear regression, expressing vaccination rate as a function of time. This assumes that vaccinations are operating at capacity (see Appendix A for a longer discussion on the reasons for this approximation, and further ways of improving this model). Having imputed the rate of new vaccinations *Π*_*s*=1…*t*_ in the days leading to the event, we turn to the estimation of the number of individuals that are likely to be susceptible. Recent reports indicate that vaccine-acquired immunity is a function of both time since vaccination and number of doses [55]. To compute the effective number of participants at risk in the event, we use a compound Poisson distribution: on each day *s* in the weeks leading to the event, the number *X* of new participants vaccinated (having either their first or second dose) is expressed as a Poisson 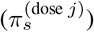), where *j* ∈ {1, 2}. Each of these newly vaccinated individuals then has a probability 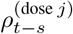 of being immune, depending on the date and dose *j* that they have received. The resulting number of immune people Z attending the event thus follows a Poisson model with rate: 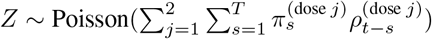. We discuss in Appendix A how this estimation can easily be modified as the vaccination rates increase and the Poisson approximation becomes no longer valid.

##### Royal Albert Hall Example

For the RAH example, we present a comparison of each quantity for three different dates (see Table 1). Of note is that the screening safety protocol is effective in more than 60% of cases that when combined with the expected infectiousness of participants and self-reporting of COVID-like symptoms, imply that 95% of infected cases are removed. We also note that prevalence is very important in determining the number of infectious cases at the event — thereby highlighting the importance of a context-aware risk calculator.

**Table 1:** Comparison of the efficiency of the screening protocol and the number of infectious participants at the event by event date. The combined effect of the screening protocol, and the natural time-dependent infectiousness of infected ticket holders mean that the number of infectious participants at the event is likely to be low, and in the order of tens in times of extremely high prevalence.

| Date of the Event | August 3rd 2020 | January 18th 2021 | March 8th 2021 |
| --- | --- | --- | --- |
| Projected incidence | 36 in 1,000,000 | 1,210 in 1,000,000 | 410 in 1,000,000 |
| Number of Infected Participants | 3.3 | 174 | 57.5 |
| Number of Infectious Participants at the Event | 0.20 | 10.5 | 5.3 |
| Percentage of Caught Cases | 71 % | 69% | 62 % |
| Number of Susceptible Participants | 4,996.7 | 4,826.0 | 4036.5 |

### Step 2. Modelling Transmission Dynamics

Having estimated the number of infectious participants at the event, the second major component of our model consists of estimating the number of transmission events during the event itself.

#### Identification of transmission mechanisms

More than a year after the start of the epidemic, the precise mechanisms by which COVID-19 is transmitted are still unclear. Aside from direct physical contact, experts continue to debate the significance of the following two main routes of infection:

##### (a) Droplet transmission

In this scenario, transmission happens through the inhalation of droplets (particles of 5 to 10 *μ*m in diameter [56]), and typically occurs when a person is in close proximity (within 1 meter) with someone who has respiratory symptoms (e.g. coughing or sneezing).

##### (b) Airborne transmission

Increasing concerns around airborne transmission have been raised by a number of experts over the past few months [57, 58]. Airborne transmission refers to the presence of the virus within droplet nuclei remaining in the air for long periods of time and with the potential to travel long distances [57] and penetrate more deeply in respiratory tracts. Airborne transmission has been estimated to be nearly 19 times more likely indoors than outdoors [59]. In the context of large public events, this transmission route thus has more diffusive power and hence could explain several super-spreader events (SSEs) [6] making it a major cause for concern [60, 61, 53, 62, 2, 57, 63, 64, 65, 66].

While droplet emission is undeniably a source of concern and a major source of transmission, simple safety precautions such as mask wearing have been shown to efficiently control this transmission source [67, 68]: it is estimated that face masks can block 80% of exhaled droplets and reduce inhaled droplets by up to 50%, and so on average reduce the transmission probability by 70% [67]. Conversely, the evidence concerning the efficiency of standard protective equipment in filtering aerosol droplets varies widely across studies probably due to “variation in experimental design and particle sizes analyzed” [67]. Airborne transmission in indoor settings can thus represent one of the main risk factors in live events, which we focus on modelling using the aerosol model proposed by Jimenez [69, 63]. The Jimenez aerosol transmission model [69, 21, 63, 70] is currently one of the only COVID-transmission models that provides enough granularity to quantify the risk associated with an event. This recognized model has been used several times in the literature over the course of the pandemic, including to allow in-class teaching at the University of Illinois at Chicago [64]. Based on the Wells-Riley model [71, 72, 73], this estimator calibrates the quanta to known transmission events, and takes into account important factors to compute a risk estimate, including event-specific (number of people, local prevalence, etc) and venue-specific variables (ventilation rate, size of the venue, UV exposure). This Wells-Riley-based model relies on the evaluation of three quantities: (a) the quanta exhalation rate, which is contingent on the activity performed and the number of infectious participants; (b) quanta concentration, which is a function of the volume of the space, the room ventilation rate, and the quanta exhalation rate; and (c) quanta inhalation rate, which is a function of the quanta concentration and breathing rate associated with the activity performed. The probability for each susceptible individual to be infected can then be written as: 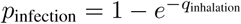 (See Appendix B for more details).

#### Modelling the uncertainty of the model

To estimate the uncertainty associated with this model, we use Monte-Carlo simulations. We simulate random input parameters (number of infectious and susceptible individuals) using the distributions and uncertainty estimates discussed in the previous section. In order to model the uncertainty associated with the Jimenez transmission model, we add a sampling step at the end of the Jimenez pipeline. This allows us to account for individual variations in infectious participants’ ability to spread the disease, and to remain consistent with the extensive literature on the heavy-tailed, Pareto nature of COVID transmission and superspreading [74, 75, 76]. For each susceptible participant, we sample their probability of infection *p*_infection_ from a Pareto distribution centered at the computed *q*_inhalation_, and with shape *θ* = 1.16 and rate 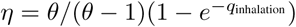. This produces a distribution centered around 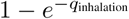, but heavily skewed to model variability in the crowd. This choice of parameters allows us to abide by the Pareto principle, according to which 80% of transmissions are due to 20% of those infected.

### Step 3. Comparison with the Null Model

To quantify the effect of the event, it is necessary to put it in context of the background rate of infections: even if the participants had not been to the event, they could have been infected elsewhere. In this null model, the number of infections is binomially distributed, such that the number infections Y is *Y* ∼ Binom(*n*_susceptible_,
π).

#### The Royal Albert Hall

We present the results for the RAH example in Table 2. This table shows in gray the values of the different quantiles of this distribution. We note the skewed distribution that we obtain is expected given the modelling of the uncertainty around inhalation rate. If the event did not occur, then on each respective date there would be an expected community transmission of 0.2 (95% prediction interval: 0.00, 1), 3.5 (0.00, 8) and 2.5 (1, 6) events on August 3^rd^ 2020, January 18^rd^ 2021 and March 8^th^ 2021, respectively. However, with the event taking place on these dates, and calculating the expected number of infectious individuals, susceptible individuals and transmission dynamics within the venue, the number of transmission events would in general increase to 0.5 (0.3, 6.8), 20 (11, 264) and 5.4 (3.0, 71) in that same order.

**Table 2:** Quantiles of the number of transmission events for the RAH concert. The grey cells provide the quantiles for the null distribution, that is, a situation in which the event does not take place but ticket holders might still get infected as a by-product of the background community prevalence.

| Date of the Event | August 3rd 2020 |  | January 18th 2021 |  | March 8th 2021 |  |
| --- | --- | --- | --- | --- | --- | --- |
| Scenario | Event | No Event | Event | No Event | Event | No Event |
| Median | 0.5 | 0.2 | 20 | 3.5 | 5.4 | 2.5 |
| 1st Percentile | 0.28 | 0 | 11 | 0 | 2.9 | 1 |
| 2.5th Percentile | 0.28 | 0 | 11 | 0 | 3.0 | 1 |
| 97.5th Percentile | 6.8 | 1 | 264 | 8 | 71.3 | 6 |
| 99th Percentile | 14.6 | 2 | 575 | 8 | 162 | 7 |

It is likely, although not inevitable, that the event will have an impact on the transmission and increase it irrespective of the level of the prevalence. However, for low levels of prevalence and higher vaccination rates, this substantially decreases. Having computed the number of expected transmission events, we can then compute several complementary metrics of interest including for example the secondary attack rate (SAR) — that is, the number of COVID cases in the participants’ community in both the null and the event model. SAR can be calculated from the predicted reproductive rate (*R*) in the regions where the ticket holders dwell. In the UK, *R* rates are updated on a weekly basis at regional levels (e.g. East Midlands, London etc) and available from the Office for National Statistics, or can be derived from the kNN modelling previously described. An opportunity for further research would be to estimate SAR within households by gathering contextual data from ticket holders. Equally, estimates of hospitalizations and deaths might be possible based on individual characteristics and comorbidities, however this is beyond the scope of the current article.

## 3. Evaluating the Effectiveness of the Screening Protocol

This risk modeling pipeline also allows comparison of different protocols and situations. For example, this pipeline highlights (a) the importance of event duration: the longer the dwell time at the event, the more at-risk the participants, and (b) the importance of wearing masks. Table 3 quantifies outcomes of holding the event on our three dates, assuming that either 0, 50 or 100% of participants are wearing masks, or varying parameters such as the density or length of the concert. Figure 4 completes that analysis by providing a visual representation the effect of these parameters on the distribution of the number of infections.

**Table 3:** Effect of different input parameters on the quantiles of the number of infections for an event at the RAH across all three dates.

| Date of the Event | August 3<sup>rd</sup>, 2020<br>(median, 95% CI) | January 18<sup>th</sup>, 2021<br>(median, 95% CI) | March 8<sup>th</sup> 2021<br>(median, 95% CI) |
| --- | --- | --- | --- |
| No mask<br>3h<br>5,000 participants | 3.4 (1.9, 46.6) | 131 (74, 1748) | 36.4 (20.3, 485) |
| 50% mask<br>3h<br>5,000 participants | 1.7 (0.9, 23.8 ) | 65 (36, 873) | 17.7 (9.9, 226.2) |
| 100% mask<br>3h<br>5,000 participants | 0.5 (0.3, 6.8) | 20 (11, 275) | 5.4 (3.0, 71.3) |
| 100% mask<br>1.5h<br>5,000 participants | 0.3 (0.1, 3.3 ) | 10 (5.6, 128) | 2.6 (1.5, 35.7) |
| 100% mask<br>3h<br>2,500 participants | 0.2 (0.1, 2.3) | 6.7 (3.8, 88.5) | 1.8 (1.0, 24.4) |

**Figure 4:**
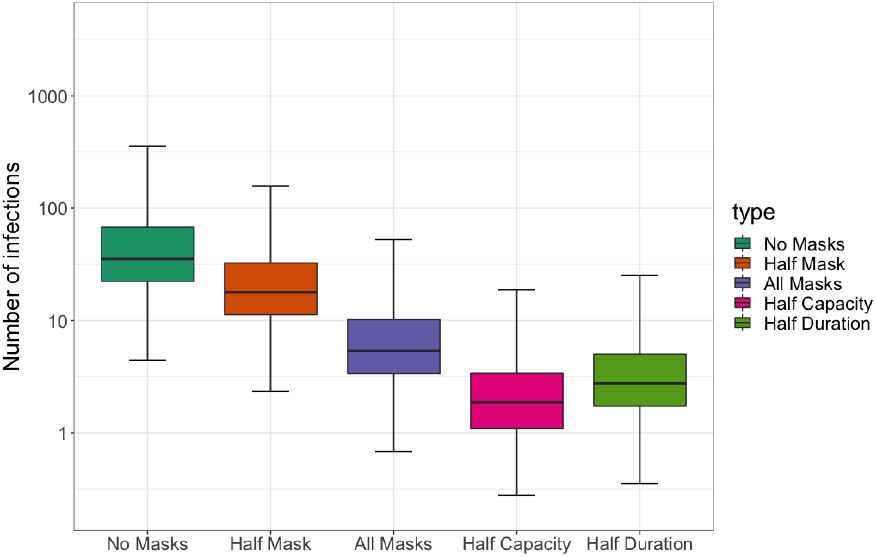
Effect of different input parameters on the distribution of the number of infections for an event at the RAH held on March 8^th^ 2021.

In addition to the aggregated risk that a live event presents, individual risk of transmission can be estimated and can be communicated to ticket holders so that they can gauge whether the risk of attending the event outweighs their desire to attend. For the first person to purchase a ticket, risk of transmission will be calculated based on their own immunity status (vaccination, regional prevalence etc) and a synthetic population based on national prevalence at that time. As more bookings are assigned to ticket holders, the reliance on the synthetic population decreases as understanding of the number of susceptible and potentially infectious individuals attending the event increases. Therefore, the confidence in the risk score increases as the event draws closer and as the proportion of tickets sold increases. This can be reflected in the updated risk scores provided to ticket holders as the event approaches. The individual risk scores can be modified based on alternative scenarios imputed into the risk algorithm. For example, for an individual not yet vaccinated, their risk could be also presented as if they had been vaccinated, offering an opportunity for the individual to appreciate how vaccination could have modified their risk. Such an approach could form the basis for behaviour change interventional studies for promoting health literacy and tackling vaccine hesitancy (see Appendix D). By working in partnership with the live events organizer, individuals that chose to opt out can be reimbursed without delay and the ticket re-sold.

## 4. Discussion

The modelling we propose is based on prevalence estimates and screening protocols to calculate the number of infectious and susceptible individuals attending the event as well as transmission dynamics at the venue to predict the number of new infections. Our paper demonstrates the value of estimating attack rates from live events so that they can be appropriately managed. We also demonstrate how individual ticket holders can receive personalized risk scores for contracting COVID-19 at the event which would, for the first time, enable genuine informed consent to be obtained. Although this methodology provides clear benefit to event organizers, local public health authorities and individual ticket holders, our approach is based on several assumptions which group in two categories: modelling assumptions and parameter sensitivity.

### Modelling Assumptions

As they combine data and tools from different sources, the computations in our pipeline rely on assumptions at three main levels:

#### (a) Predicting COVID-19 prevalence

To predict future COVID-19 incidence, we chose a kNN approach as it yields a more robust prediction and better uncertainty quantification than most existing parametric methods. One of the downsides of this approach is that it might not generalize very well to entirely novel behaviours or viral variants − in which case well-parameterized methods may outperform our approach as knowledge of transmission, vaccination and other relevant model parameters continues to improve. While prevalence predictions are important for event planners and attendees alike, on the day of the event the more important metric is whether official case rates reflect actual cases, i.e. the ascertainment rate. Historically, this rate has been low due to limited testing facilities, and our method to determine ascertainment using cases, deaths and infection-fatality rates reflects this, but also indicates that ascertainment may exceed 100% in times of widespread testing and low prevalence. It was beyond the scope of this paper to further investigate ascertainment but we expect that future research will clarify the impact of different test types, their false negative and positive rates, and their frequency of use in determining the ascertainment rate.

#### (b) Assessing the efficiency of the screening protocol

Our modelling framework assumes that events will screen participants with COVID-19 tests, such as virtually-witnessed lateral flow antigen tests. Assessing the efficiency of this screening step requires the estimation of (a) the sensitivity of the test, (b) the probability of having symptoms, and (c) the probability of being infectious − all of these quantities being a function of days since infection. Our estimation of each of these quantities is based on published data - with the exception of the probability of Symptom Check failure (i.e, the probability that a participant lies about their symptoms to get in). By default, we select this probability to be 50%, a choice that will be improved upon as the CAPACITY and other similar studies gather behavioural data. However, as shown in Appendix C, this factor has a relatively minor impact on the outcome of the model compared to the uncertainty of the other inputs. Of potentially greater concern is our assumption that the probability of testing negative 2 days before the event is independent (conditionally on time since infection) of a participant’s infectiousness during the event. A potential avenue for improvement could consist of determining both test sensitivity and infectiousness as a function of viral load, and estimating the joint probability of the viral load 2 days apart. However, the data required for this approach is − to the best of our knowledge − still lacking, and given the variability of the viral load or PCR Ct behaviour, this conditional independence assumption seemed a reasonable first-order approximation.

#### (c) Transmission at the event

The airborne transmission model that we use relies on an homogeneous (well mixed) air hypothesis. While several other models have been proposed (either breaking the room into compartments or using a distance index) to counter (disprove or annul) this hypothesis, we highlight (following the discussion by Jimenez [69]) that this is a first order approximation: some participants will have more risk and others less, so that at low quanta concentration, this effect will be averaged out. At very high concentration, the model will likely under-estimate the number of infections, but given the efficiency of the screening protocol and density limitations, we do not expect this scenario to be common. Finally, we note that our model is not tied down to any specific transmission mechanism, and as our knowledge of COVID transmission improves, we can refine and supplant the transmission dynamics with a superior alternative or another model that is deemed more suitable.

### Parameter Sensitivity

While we try to limit the number of input parameters in our pipeline, the sensitivity of the estimates to these inputs (namely, the mask efficiency and population of interest) has to be studied. We refer the reader to Appendix C for a quantitative sensitivity analysis and highlight our conclusions here. In terms of the model parameters, the greatest unknown consists in determining the efficiency of masks and protective equipment - the latter having been shown to vary depending on the mask type and activity. However, we hope to make use of the growing body of literature on the topic to update and refine this important factor. Secondly, our prediction framework assumes that participants at the event have the same probability of infection and vaccination as their regional average. However this might not be the case as participation in the event may be an incentive to get vaccinated, or conversely might select for less cautious sub-populations. The importance of this sampling frame assumption nonetheless decreases as participants’ vaccination status and behavioural data from the CAPACITY study will result in more precise estimates.

## 5. Conclusion

A nuanced, data-driven system is required to assess risk at each event informed by the characteristics of all ticket holders and the background risk of transmission concurrent to the event, so that proportionate and specific action can be taken by event organizers and public health authorities. We have detailed our attempt to create such a system and have outlined its predictions and limitations. Our end-to-end risk model is provided in the form of an R-shiny interface. At times of high prevalence, this type of system will ensure events likely to increase transmission can be halted. At times of low prevalence this will ensure events can potentially continue to operate. Learning to live with SARS-CoV-2 will be about implementing systems that support hyper-local, data driven decisions so that far-reaching and highly damaging sector-specific lockdowns can be avoided as much as possible.

## Data Availability

The links to all datasets used in this study are explicitly provided in the paper.

## Acknowledgments

The work of TE has been supported by the Estonian Research Council Grant PRG1291. MH and AEL are supported in part by the NW London NIHR Applied Research Collaboration. Imperial College London is grateful for support from the NW London NIHR Applied Research Collaboration and the Imperial NIHR Biomedical Research Centre. JK is currently Director of Health Optimisation at the Center for Health and Human Performance (London, UK), as well the co-founder and Medical Director of CERTIFIC.

## Disclaimer

The views expressed in this publication are those of the authors and not necessarily those of the NIHR or the Department of Health and Social Care at Imperial College London, nor neither of any of the authors’ corresponding institutions (University of Chicago, Carnegie Institution for Science, and University of Tartu).

## A. Appendix: Prediction

As explained in the introduction, the aim of this paper is to develop a context-aware risk model: to be informative, the estimates of the risk that the model outputs must informed by (a) the prevalence at the time of the event, (b) the ticket holders’ vaccination status, and (c) the screening protocol employed by the event management to reduce transmission risk. In this setting, the CAPACITY protocol serves as a case in point and a motivation to our paper. Central to the study is the estimation of the risk associated with the event on two different horizons:

### Horizon 1: Weeks prior to the event

In this setting, the goal is to predict ahead of time the risk associated with the event, and help organizers and ticket holders alike to plan ahead and decide whether or not they deem the risk associated with the event acceptable. This step requires the prediction of both the prevalence of the disease and the vaccination status of the crowd several weeks in advance.

### Horizon 2: A few days before the event

The purpose of the risk estimation is to evaluate—with more certainty the admissibility of the risk associated with the event. In this step, the algorithm can rely on ticket holders’ reported vaccination status, as well as the most recent incidence rates to compute the risk.

Thus, whilst not crucial for Horizon 2 (in the last few days leading up to the event), the prediction of the crowd’s vaccination status as well as the incidence rate are a major component of Horizon 1— thereby calling for prediction methods that both provide accurate estimates and a correct evaluation of their associated uncertainty. In this appendix, we focus on providing more details into the different predictive components that we use to estimate the risk ahead of time (Horizon 1). These consist of the following three main steps, which we subsequently describe in greater details:

- Step a: The prediction of the number of newly infected individuals who are ticket holders.
- Step b: The prediction of the number of infected participants that will escape the screening protocol.
- Step c: The prediction of the number of vulnerable ticket holders at the time of the event (i.e, participants that are not immune).

### Step a. Prediction of new cases through a k-Nearest Neighbour (k-NN) Approach

The first step in our pipeline consists of the estimation of the daily incidence rate in the days leading to the event, which, in the main text of this paper, we suggested solving using a *k*-nearest neighbour approach with k=100. Indeed, this choice of k=100 neighbours allows us to have sufficient information to evaluate the percentiles of the distribution of the prediction, whilst retaining a sufficient amount of similarity with the original trajectory. The algorithm is described in details in Algorithm 1.

#### Algorithm 1 Prediction of the epidemic curve using k-NN

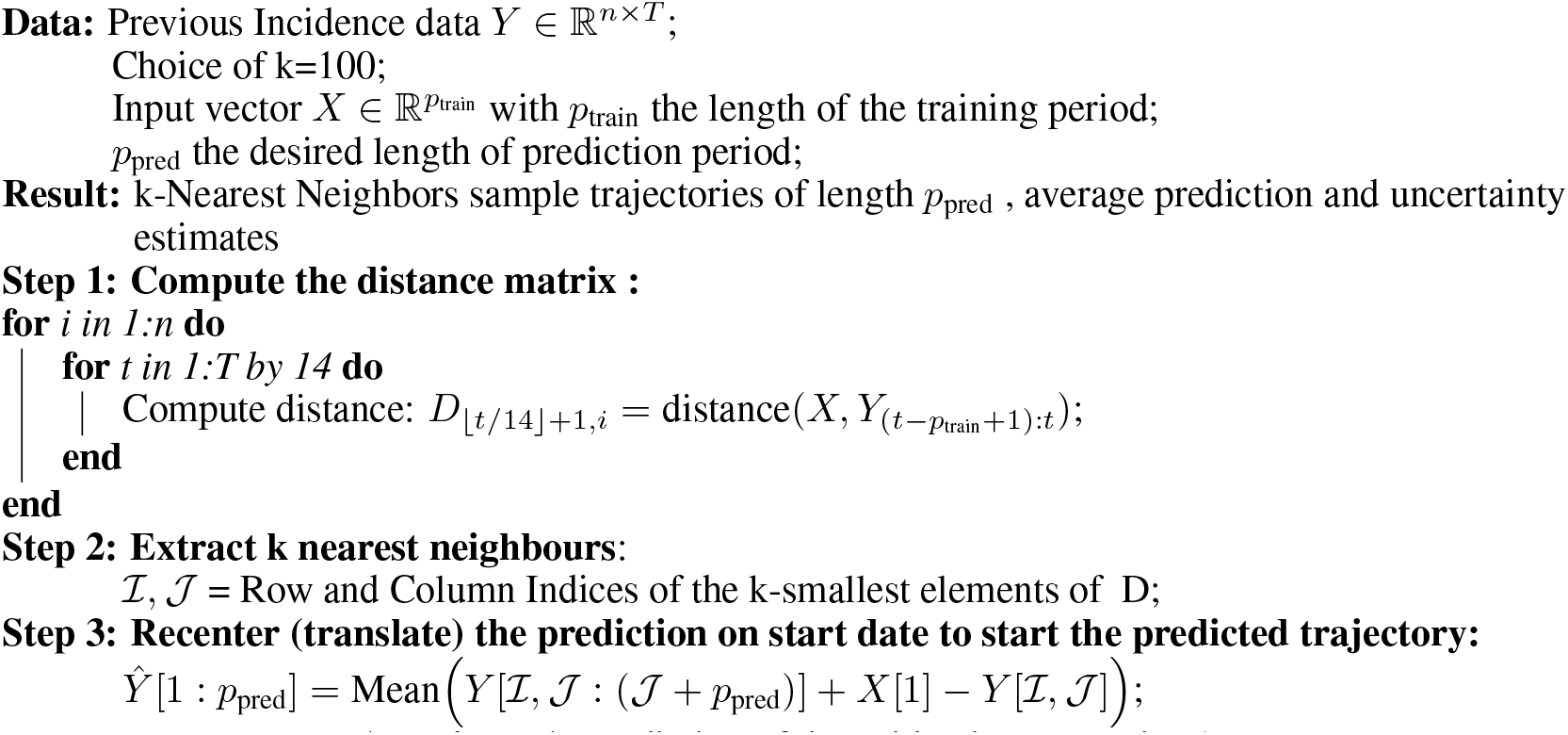

#### Motivation for the k-NN approach

COVID-19 prediction is undoubtedly an involved task — as denoted by the impressive amount of literature published on the topic [34, 35, 36, 37, 38, 39, 40, 41, 42, 43, 44]. Yet, as emphasized in the main text, many of these methods rely on a parametrization of the problem (Exponential growth, SEIR model, etc. [45, 46]) that require input parameters (eg, the reproductive number) that are both unknown and non-stationary. Indeed, as the number of cases rises, policy makers are bound to adapt their policies to limit the spread of the virus. Reciprocally, as community prevalence levels drop, strict lockdown measures and stay-at-home orders are bound to be lifted—thus impacting the transmission modalities and likelihood of propagation of the virus. Incorporating both uncertainty and non-stationarity in predictions is thus a challenging task. Our solution to this problem relies on using a k-Nearest Neighbour approach (k-NN). To predict the trajectory of the epidemic in the next *p*_pred_ days, we use daily incidence (per hundred people) data from all countries around the world at any time during the epidemic, and look for the 100 curves that best match our country/region of interest in the past 28 days. This comparison is done using a simple 𝓁_2_ distance (we discuss in the following paragraph other choices of distances). The choice of 28 days is motivated by the fact that we need sufficient data to find trajectories with similar behaviours, whilst remaining sufficiently local for the comparison to be valid. Having found these 100 closest neighbours and translated them adequately to make them start at the observed value for the trajectory of interest on day 0, we use these 100 closest neighbours’ observed trajectories in the next *p*_pred_ days to deduce an estimate for our future trajectory and an associated prediction interval. The underlying assumption here is that these observed 100-NN curves contain information both on the reproductive number and propensity of the epidemic to grow, but also on policy decisions made as a result of rising (or declining) prevalence numbers — thus making them an appealing non-parametric candidate for incidence modelling.

#### Performance and Benchmarking of the k-NN approach

To provide more substantial ground for our proposed kNN approach, we studied its performance compared to more traditional benchmarks using exponential growth model and Attack rate models [46, 45], commonly used in the literature and implemented in the R package R0 [77]. We compare a mix of classical projection methods, as well as k-Nearest Neighbour methods using various distances:

- The Attack Rate method [45], using the R package R0 [77], Incidence [78] and Projections[79],
- The Exponential Growth model [46] (using the same R packages),
- Our k-NN method, computing the distance between the training trajectory and the dataset using the 𝓁_2_ distance (we will denote this method by the “Sum of Squares”)
- Our k-NN method using a weighted 𝓁_2_ distance, in which the weights of observations are inversely proportional to the time since the origin of prediction (ie: for each day 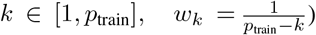. This weighting of the observations puts more emphasis on more recent observations, so that trajectories that share a higher degree of similarity in their recent behaviours will be favoured by the algorithm. We will denote this method by “Weighted Sum of Squares”.
- Our k-NN method, computing the distance between the training trajectory and the dataset using a correlation measure between trajectories.
- Our k-NN method, computing the distance between the training trajectory and the dataset using a correlation-based distance, whilst giving more weight to recent observations (in a similar manner than for the “Weighted Sum of Squares” approach).

To compare these methods, we use data from “Our World in Data": for each country, we evaluate the performance of each method for predictions from June 1st 2020 to February 14th 2021. Every 28 days, we recreate a prediction scenario, in which we use 4 weeks of observations for training, and predict the daily incidence for the next four weeks. The purpose of this experiment is to assess and compare across methods (a) the mean square error as a measure of the methods’ prediction accuracy, and (b) the coverage (percentage of times that the confidence interval covers the true observations), in order to establish which method allows to correctly estimate the uncertainty and risk. The results across countries and periods are provided in Figure 5a and the coverage is shown in Figure 5b. We see that the k-NN method achieves comparable MSE to the classical projection methods implemented in R. Yet, the k-NN method also achieves a coverage of more than 90% (and close to the nominal 95% that it targets), and the prediction interval that it provides is thus more reliable than that of other methods. Interestingly, we also note that the weighted and unweighted “Sum of Squares” k-NN achieve roughly the same performance. For the sake of simplicity, we thus use its unweighted counterpart.

**Figure 5:**
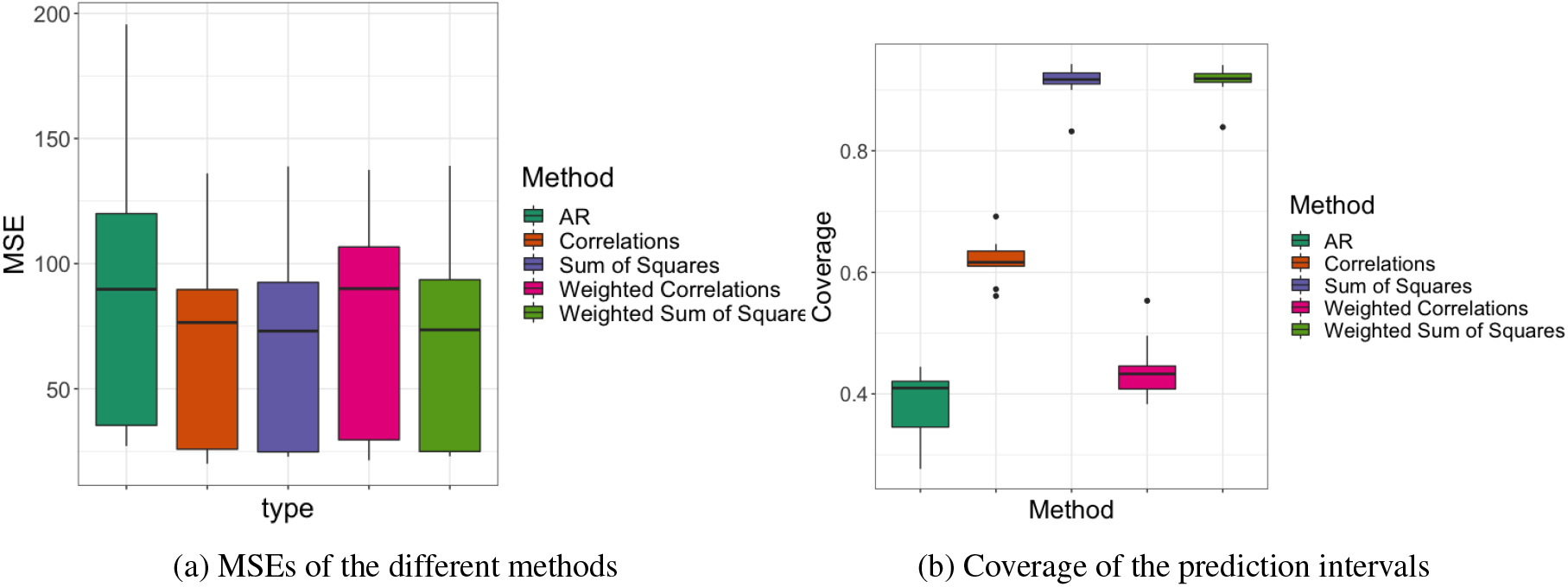
Performance of the different methods for predicting the epidemics trajectories (all countries around the world) (a) Ascertainment rate for the United Kingdom. The y-(b) Vaccination rate: comparison of the actual rates, and the axis denotes the ratio of detected cases to cases predicted ones predicted by a linear model for the United Kingdom. by multiplying a 3-week shifted death rate by a calculated The vaccination rates (both first and second dose) seem to infection-fatality rate for the UK. When the values are less be well approximated by a linear regression model (using than one it suggests that many of the actual cases are not time as a covariate), with an associated *R*^2^ of 0.92. As being detected, and when the values are greater than one time progresses, and vaccination rates increase, this linear it suggests that either a substantial proportion of cases are fit might start becoming less accurate, as problems of vacci-false positives or that survival from COVID infection has nation hesitancy or difficulty to access might induce vaccine increased from baseline predictions, due to better treatments centers to operate under capacity. or a greater bias towards infection of individuals at lower risk of death

### Under-ascertainment bias

Having predicted the daily incidence rate in the weeks leading to the events, to correctly estimate the number of ticket holders that are likely to be contaminated, it is important to correct this prevalence estimate for any under-reporting bias. The under-ascertainment bias refers to the fact that the reported COVID cases are in fact an under-estimation of the actual number of cases, due to either asymptomatic cases or limited testing capacities. In order to compute the appropriate correction, we use age-stratified estimates of the Infection-Fatality rate ^4^. The method provided at this link has the advantage of computing robust IFR estimates by leveraging data from countries around the world, and adjusting for their demographic makeup. The actual number of cases is then computed as:

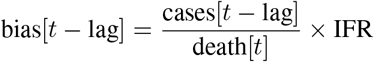

Indeed, since deaths are offset compared to the incidence rate, reported new cases must be compared to deaths roughly three weeks later (according to CDC reports). The under-ascertainment in the case of Britain is plotted in Figure 6a.

**Figure 6:**
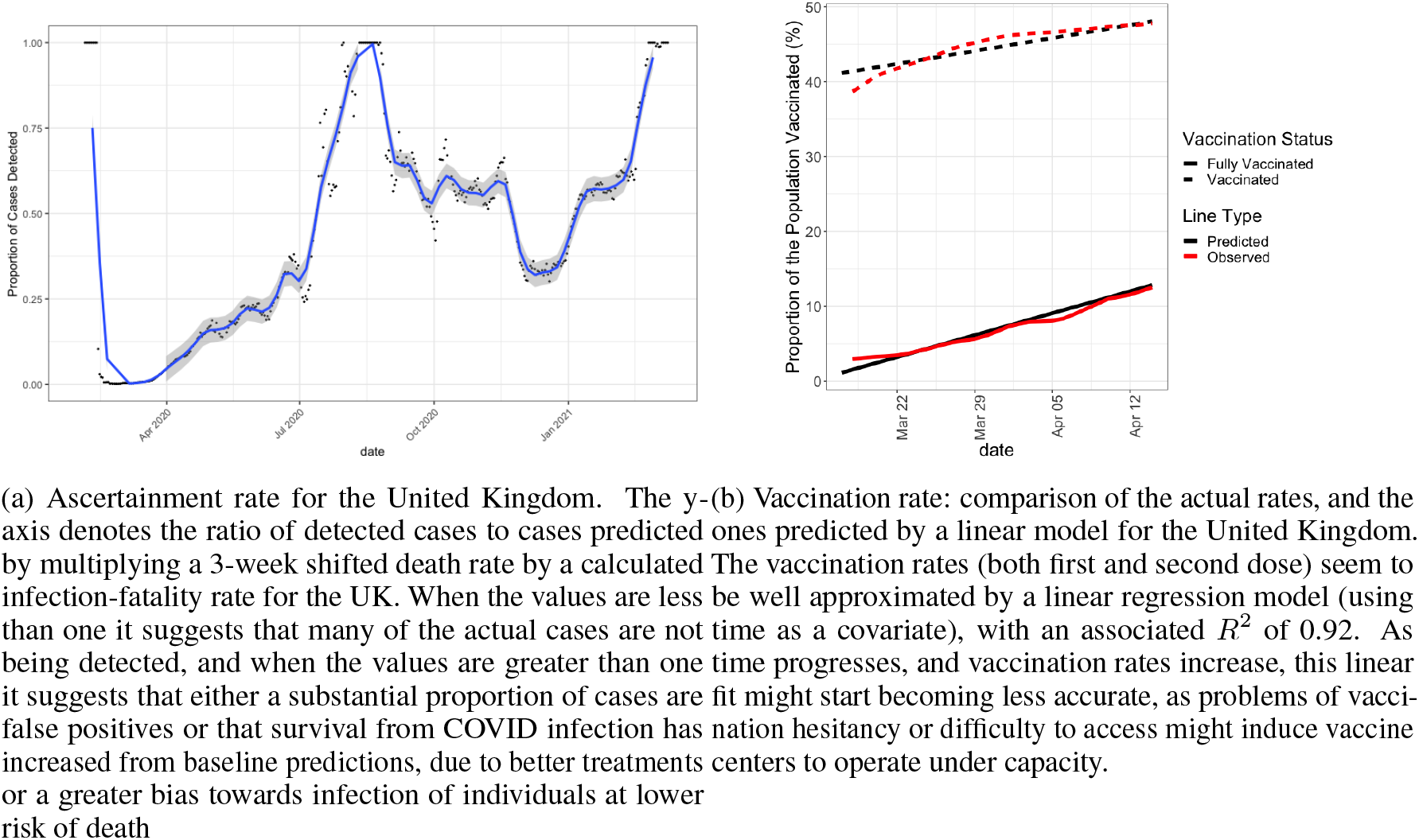
Under-ascertainment rate (left) and vaccination rate (right) in the United Kingdom.

### Step b. Estimating the Number of People who will escape the screening protocol

For the sake of clarity and to make this appendix self contained, we repeat here the discussion of the screening protocol provided in the main text, but provide additional detail on the estimation procedure. For an infectious individual to attend the event in spite of the CAPACITY study’ screening protocol, they must (a) have no COVID-like symptoms or fail to report them on the morning of the event, (b) receive a (false) negative result during antigen testing D = 2 days prior to the event, and (c) be contagious (rather than simply infected) at the time of the event.

#### (i) Symptoms-Check Failure

Indeed, one of the main challenges associated with the COVID-19 crisis is the number of asymptomatic cases - that is, infected individuals that do not express symptoms and are thus unaware of their potential infectiousness. This group encompasses people that are either pre-symptomatic or completely asymptomatic during the course of their illness − the latter are estimated to represent roughly 25% of all cases [47]. To account for this temporal dependency, we use estimates of the incubation period (defined as the number of days between infection and symptom onset) from McAloon et al. [48] and data on symptoms duration from van Kampen et al. [49] to estimate the probability for a ticket holder infected *s* days before the event to exhibit symptoms on the day of the event. We rely on simulations to estimate this probability distribution, finding estimates of the time to symptom onset by randomly generating an incubation period using data from McAloon et al. [48], and sampling a symptom duration from van Kampen et al. [49]. The resulting density plot is displayed in red in Figure 3a.

#### (ii) Antigen test failure

The sensitivity of COVID tests depends heavily on the time since infection, and whether these are the gold-standard PCR or Lateral Flow Antigen Assays [50]. Moreover, studies have shown that LFA tests have much lower sensitivity on asymptomatic individuals than symptomatic: in particular, according to a recent CDC report [51], Rapid Antigen testing has 80% sensitivity on symptomatic individuals, but only 40% sensitivity on asymptomatic individuals. Coupling the sensitivity estimates [50, 51] with the distribution of incubation period and estimated percentage of asymptomatic cases [48, 47], for each individual infected at day *k* taking an antigen test *D* days before the event, the probability of getting through the filtering protocol is thus given by the formula:

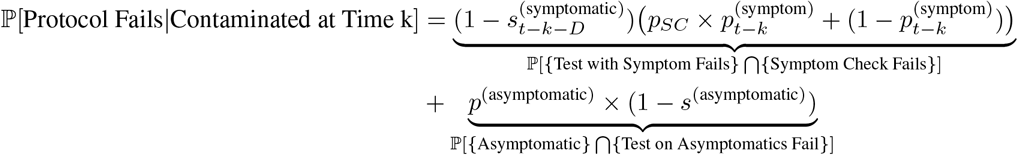

where 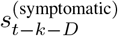 and *s*^(asymptomatic)^ are respectively the sensitivities of the test taken *D* days before the event for a symptomatic participant infected *t − k* days before the event and an asymptomatic individual. The parameter 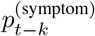 denotes the probability for a symptomatic individual to exhibit symptoms *t − k* days after infection, whereas *p*^(asymptomatic)^ is the probability of being asymptomatic. Finally, the variable *p*_*SC*_ denotes the probability of the Symptoms Check failing—namely, that the participant does not want to report their symptom. Currently, this probability is set by default in our model to 50%, and we provide in our interactive dashboard the option to choose other levels. As the CAPACITY study gathers more behavioural data on the participants, we hope to improve this estimate. However, we study the sensitivity of our analysis to the choice of this variable in Appendix C, and show that in view of the total uncertainty surrounding other parameters in the model, the choice of this parameter does not severely affect the robustness of the results.

Figure 3b in the main text shows the probability of the failure of the screening protocol as a function of days after infection. This curve was also simulated by sampling: we model the uncertainty in the sensitivity through a set of Monte Carlo simulations, in which, for each simulation: (a) we sample a random sensitivity from a beta distribution, with parameters chosen to match the uncertainty intervals provided in [50], and (b) associate these random sensitivity to the probability of having symptoms and failing to report them. The shaded areas in Figure 3b denote the uncertainty around this estimate due to the variability of the incubation time.

#### (iii) Estimating Infectiousness

Infectiousness is a function of time since infection. Many articles in the literature have in particular estimated infectiousness to be at its peak within the first five days after symptom onset. However, very few reports provide an in-depth description of infectiousness as a function of time since infection. To this end, in this paper, we combine data from multiple sources. In particular, we rely on the data from Singanayagam et al [52]. Indeed, in this article, the authors study infectiousness as a function of time since symptom onset which they estimate by looking at the percentage of viable cultures that they can obtain from samples collected at various intervals before and after symptom onset. Since our goal is to consider infectiousness as a function of time since infection (rather than symptom onset), we combine this data with the estimated distribution of incubation length (duration between the date of infection, and date of symptom onset). One of the main issues in converting the data from [52] lies in the long tails of this distribution, which extrapolates from the data and allows samples to be highly contagious up to 10 days before infection, Since Singanayagam et al [52]have very few cases past 4 days prior since infection, we threshold infectiousness to 0 to be consistent with the estimates by He et al [53]. We compound the distribution of incubation and infectiousness by a probability of the incubation length, yielding a probability of infectiousness *s* days after infection such that:

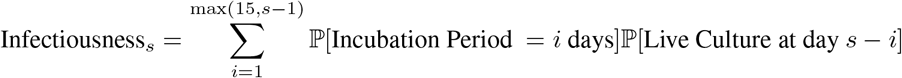

### Step c: Estimating the number of people at risk

Finally, the last quantity that we need to impute is the number of people at risk during the event. As described in the main text, this requires a knowledge of the participants’ COVID immunization status, i.e, has the participant already had COVID in the previous year and/or has the participant been vaccinated. This immunization status could be imputed through the combination of information regarding vaccination status as well as additional questions (previous positive test for COVID, symptoms, etc, combined in a model such as in [54]). However, for the sake of simplicity, we only consider here the vaccination status of the participants - thus leaving out the proportion of the population that has had COVID but has not been vaccinated yet. This induces a risk estimate that is biased upward—that is, we do not account for the immunity naturally gained by ticket holders through COVID infection—, and as such, is more conservative. When imputing the risk for Horizon 1 (a few weeks before the event, and without any ticket holders’ information), we impute the event’s crowd immunity level using linear regression. In other words, we assume that the number of new vaccinations (first and second dose) grows linearly each day, which amounts to assuming that vaccinations are operating at capacity. Figure 7 shows a plot of the cumulative number of first and second doses in the UK as a function of time, highlighting a good fit between the linear model and the actual observations. We note though that as vaccination levels are increasing, the linear model will potentially have to be modified: after a certain proportion of the population has been vaccinated, vaccinations could stop operating at capacity since the remainder of the population could either have difficulties in gaining access to the vaccine, or could be opposed to the vaccine altogether. However, at the time of writing, the linear fit seems to be a good fit. Having imputed the rate of new vaccinations *π*_*s*=1 *t*_ in the days leading to the event, we turn to the estimation of the number of individuals that are likely to be susceptible. Recent reports indicate that vaccine-acquired immunity is a function of both time since vaccination and number of doses [55]. To compute the effective number of participants at risk in the event, we use a compound Poisson distribution: on each day *s* in the weeks leading to the event, the number *X* of new participants vaccinated (having either their first or second dose) is expressed as a Poisson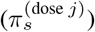, where *j* ∈ 1, 2. Each of these newly vaccinated individuals then has a probability 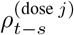 of being immune, depending on the date and dose *j* that they have received. The resulting number of immune people Z attending the event thus follows a Poisson model with rate: 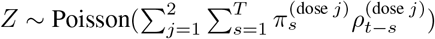

**Figure 7:**
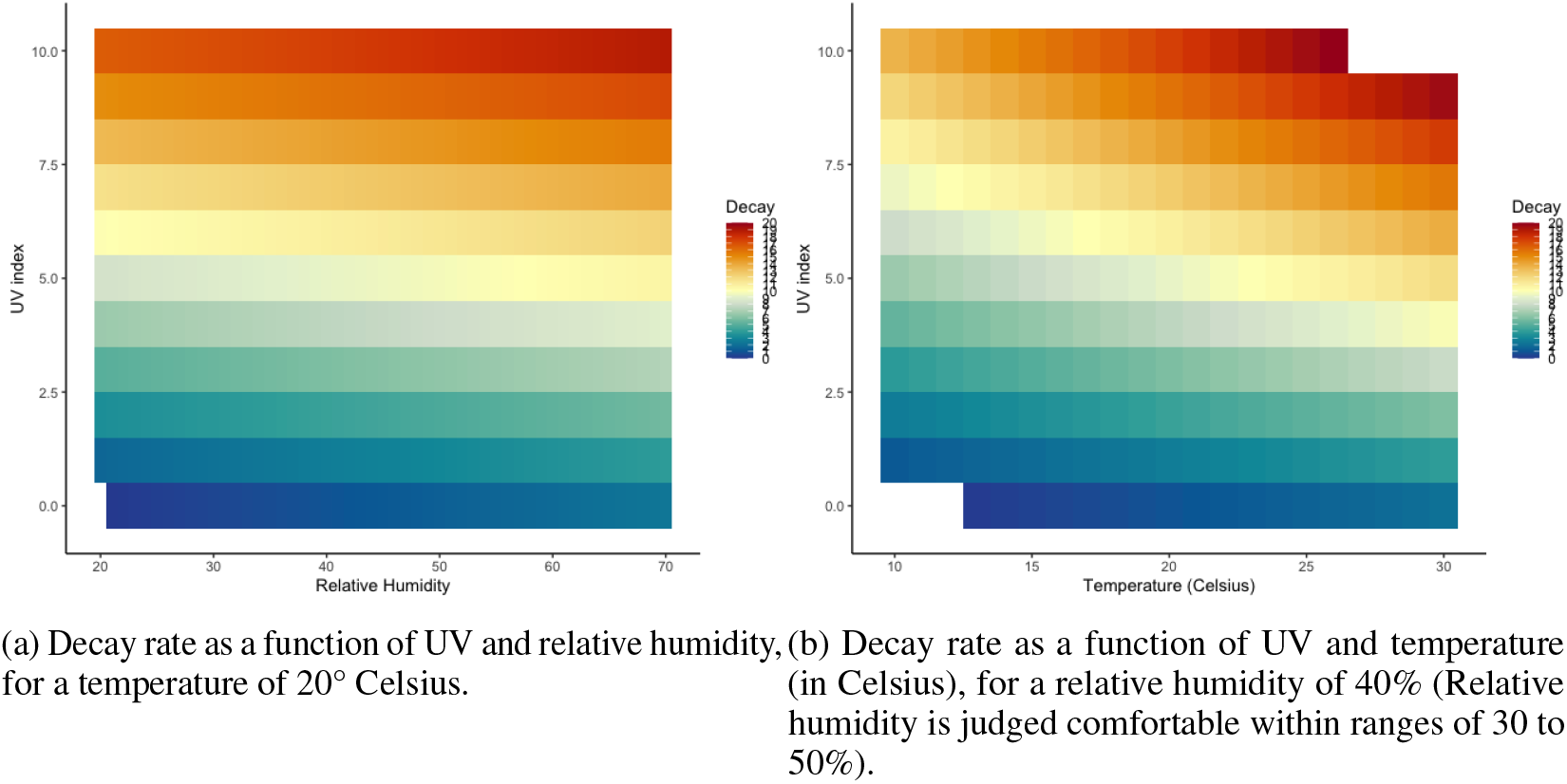
Decay rate of the virus, reproduced from [86, 87], and as a function of humidity, UV index, as well as temperature. NA values appear as transparent tiles.

However, as the number of vaccination increases, we expect the probability of the participants being immune to increase. In this case, we simply replace the Poisson binomial with a binomial, for various values of 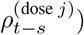 to allow the uncertainty around the immunity to percolate through the model.

## B: Appendix: Transmission

In this appendix, we describe in greater details the model and assumptions made by the Jimenez aerosol transmission model proposed in [69, 63] and used to model transmission dynamics in the main text.

### The issue of COVID transmission

As discussed in the main text, the precise mechanisms by which COVID-19 is transmitted are still unclear. Aside from direct physical contact, experts continue to debate the significance of the following two main routes of infection:

#### (a) Droplet transmission

In this scenario, transmission happens through the inhalation of droplets (particles of 5 to 10 *μ*m in diameter [56]), and typically occurs when a person is in close proximity (within 1 meter) with someone who has respiratory symptoms (e.g. coughing or sneezing). In the context of live events, modelling this specific transmission route involves (a) modelling the distribution of the number of close contacts between infectious and susceptible ticket holders during the event and (b) modelling the transmission probability for each close contact. The latter is a function of the proximity between participants and the amount of time spent in the vicinity of the infectious individual.

#### (b) Airborne transmission

Increasing concerns around airborne transmission have been raised by a number of experts over the past few months [57, 58]. Airborne transmission refers to the presence of the virus within droplet nuclei remaining in the air for long periods of time and with the potential to travel long distances [57] and penetrate more deeply in respiratory tracts. Airborne transmission has been estimated to be nearly 19 times more likely indoors than outdoors [59]. In the context of large public events, this transmission route thus has more diffusive power and hence could explain several super-spreader events (SSEs) [6] making it a major cause for concern [60, 61, 53, 62, 2, 57, 63, 64, 65, 66]. In this setting, infection risk is typically modelled using one of two distinct models: Wells-Riley equations and the dose-response model [80, 73]. First introduced by Riley in 1978 in a study of measles outbreak [71], the Wells-Riley equations are based on the concept of a hypothetical infectious unit called a “quantum of infection” [72], defined as the number of infectious airborne particles required to infect a person. Quanta aim to capture in a single parameter the rate of emission of viral particles in exhaled breath, the infectivity of the viruses upon emission, the particle size distribution of the emissions, the deposition efficiency and deposition location in the respiratory tract of the susceptible person, as well as the probability that deposition leads to infection. The dose-response model aims to describe more directly the effect on organisms from the exposure to different doses of chemicals, drugs, radiation, biological agents, or other stressors — and more recently to assess the infection risk of airborne transmissible pathogens. The review by Sze To et al [73] provides an in-depth comparison of the two models. In the context of COVID transmission, one of the main limitations of the dose-response model is that it relies on infectious dose data to derive the dose-response relationship. By contrast, many of the parameters in the Wells-Riley equation can be approximated and has thus been favored by many experts in the field [63, 70].

### Choice of Transmission Route for this Model

While droplet emission is undeniably a source of concern and a major source of transmission, simple safety precautions such as mask wearing have been shown to efficiently control this transmission source [67, 68]: it is estimated that face masks can block 80% of exhaled droplets and reduce inhaled droplets by up to 50%, and so on average reduce the transmission probability by 70%. Conversely, the evidence concerning the efficiency of standard protective equipment in filtering aerosol droplets varies widely across studies probably due to “variation in experimental design and particle sizes analyzed” [67]. Airborne transmission in indoor settings can thus represent one of the main risk factors in live events, which we focus on modelling using the aerosol model proposed by Jimenez [69, 63]. The Jimenez aerosol transmission model [69, 81, 63] is indeed currently one of the only COVID-transmission models that provides enough granularity to quantify the risk associated with an event. This recognized model has been used several times in the literature over the course of the pandemic, including to allow in-class teaching at the University of Illinois at Chicago [64]. Based on the Wells-Riley model [72], this estimator calibrates the quanta to known transmission events, and takes into account important factors to compute a risk estimate, including event-specific (number of people, local prevalence, etc) and venue-specific variables (ventilation rate, size of the venue, UV exposure).

A core principle behind the Wells-Riley model is the notion of “quantum of infection”. Exposure to one quantum of infection gives an average probability of 63% = 1 − *e*^−1^ of becoming infected (essentially an infectious dose 63%, ID63) [82]. The crux of the Wells-Riley equation consist in its modelling of the probability of infection *P*_*I*_ as a function of the ventilation, inhalation rates and quanta generation rates:

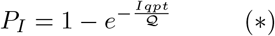

where *n*_*I*_ is the number of infectors, *p* is the pulmonary ventilation rate of a person, *q* is the quanta generation rate, *t* is the exposure time interval, and is the room ventilation rate with clean air. Note that this equation is not dimensionless. As explained by Rudnick and Milton [82], *q* represents the generation rate of infectious doses, not organisms or infectious particles; it is the average infectious source strength of infected individual. Thus, the exponential form of the probability equation reflects the probability of a susceptible person in the room inhaling at least one quanta, based on a Poisson distribution of the number of discrete quanta inhaled by a susceptible person present in the space, given a certain aerosol quanta concentration in the room and an inhalation time.

One of the advantages of the Wells-Riley model is that many extensions have been studied, allowing the incorporation of additional influencing factors. In particular, the effect of respiratory protection can be considered by multiplying the term in the exponential by a fraction [83, 84, 85]:

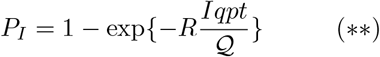

where *R* is a number between 0 and 1 representing the fraction of particle penetration of the respiratory protection (it is in particular equal to 1 when no respirator is used). Other variables, such as the ultra-violet irradiation, particle filtration have been taken into account in the Wells-Riley equation through the equation:

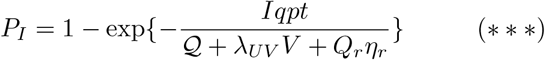

where *λ*_*UV*_ is rate coefficient of inactivation by ultraviolet irradiation, *V* is the room volume, *Q*_*r*_ is the flow rate to the filter, and *η*_*r*_ is the filtration efficiency.

As suggested by by Jimenez et al [69, 63], instead of modelling each of parameters in the Wells Riley equation explicitly, we can use the concept of quanta, and calibrate the emission rate to known outbreaks of the disease. We base the following description of the model as detailed by Jimenez et al [69, 63]. This model relies on the computation of three main components:

#### 1. The Quanta Emission Rate

The quanta emission rate can be interpreted as the number of quanta emitted by unit of time by a single infectious participant. It can be modelled as:

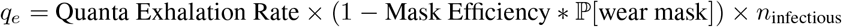

#### 2. The Quanta Concentration Rate

The quanta concentration rate *q*_*c*_ is computed as:

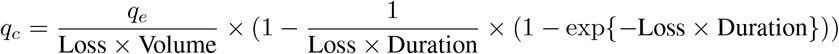

where *q*_*e*_ is the quanta emission rate and *n*_infectious_ is the number of infectious people at the event. The loss corresponds to the first order loss:

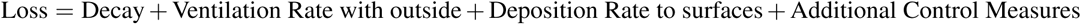

The term “Decay” corresponds here to the decay rate of the virus, and is a function of the UV, temperature and relative humidity of the event. The Decay rate per hour is computed according to the following formula [86, 87] for which an open-source calculator is available online ^5^:

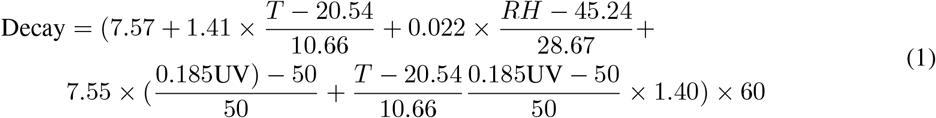

where T is the temperature and UV is the UV index. The tool is valid for the following ranges of conditions: 10 to 30°C (50-86°F), 20-70% relative humidity, and UV indices of 0-10. For live events in an indoor setting, the UV parameter should be set to 0.

We have checked that our computations are aligned with the figures provided in the references.

#### 3. The Quanta Inhalation Rate

The quanta inhalation rate is computed as:

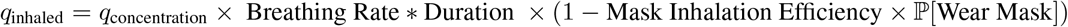

Values for the Mask Inhalation Efficiency, as well as for the breathing rate (which varies by activity) can be found at the bottom of this page. For the sake of completeness and to make this manuscript self-contained, we have included the tables suggested by Jimenez [69] in Appendix E (Figures 12, 13, and 14), and refer to the sources he suggests for a more fine-grain estimate of what these should be^6^.

### Ventilation Rates

The ventilation rate per person is computed as:

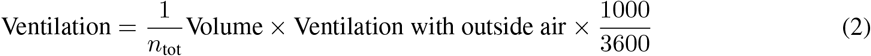

where *n*_tot_ is the total number of participants, and the ventilation rate is measured in *L*^−1^*/s* and depends on the activity. Again, for the sake of completeness, we have included in Appendix E (Figure 15)the table from the AHRAE (American Society of Heating, Refrigerating and Air-Conditioning Engineers) listing all the activities as well as their corresponding ventilation rates ^7^ that were suggested by Jimenez [69] to perform the computations.

## C. Appendix: Sensitivity Analysis

Whilst we have based our estimates of the different parameters used in this model on the literature, and tried to incorporate estimates of their uncertainty to try and correctly estimate our confidence in the output of the model, this is nevertheless contingent on several choices on (a) the probability that a ticket holder will lie and fail to report symptoms on the day of the event in order to get into the event, and (b) the efficiency of the masks, and input parameters in the room.

### (a) Probability of lying

As explained in the introduction, the risk model that we aim to develop has to be context-aware. That is, the estimates of the risk that the model should output should depend (a) on the prevalence at the time of the event and (b) on the ticket holder’s vaccination status. The only input that requires to be determined is the propensity of people to lie if they have symptoms. This is a priori a difficult parameter to estimate, which would be required to be informed by sociology studies. As the CAPACITY study proceeds, this is in particular one of the parameters that we hope to be able to inform better. However, in the absence of information as to what value of that parameter should be set, we propose here a sensitivity analysis to show that the model is in fact relatively weakly sensitive to the choice of this particular parameter. We show the different infectiousness curves corresponding to different values of the parameter *p*_lie_ in Fig. 8 and 9. As shown in Fig. 8, this probability of lying impacts the value of the maximum probability of infectiousness at the event: this value is maximal at 4 days before the event, with a value of 42.1% if the participants never lie, 48% if these participants lie half of the time, and 53% if they always lie. This represents a 25.8% increase in probability from a scenario where participants are considered as completely trustworthy to one where these participants are considered as unreliable. Table 4 quantifies the impact of this parameter in the case of the Royal Albert Hall in order to assess the sensitivity of the entire pipeline to this particular choice of parameter in two situations: low prevalence (August 3rd 2020) and high prevalence (January 18th 2021). As denoted in this table, the impact of this parameter is small in situations where the prevalence is already small. More substantial deviations occur when the prevalence increases, and such deviations are especially important in the tails (here, the median increases by 6 cases (35%) while the 97.5th prediction interval increased by 29% (69 cases) in situations of high prevalence between *p*_lie_ = 0 and *p*_lie_ = 1. While the relative increase (29%) is important, the absolute difference (69 cases) is small compared to the uncertainty in the prevalence (1245 (sd 375) cases).

**Table 4:** Quantitative comparison of the sensitivity of the results (number of transmissions at the event) as a function of *p*_*SC*_. While While the absolute difference between the scenarios remains reasonable for the median, this difference becomes particularly important in the tails of the distribution (97.5th and 99th quantiles).

|  | August 3rd, 2020 |  |  | January 18th, 2021 |  |  |
| --- | --- | --- | --- | --- | --- | --- |
|  | Never Lies | 50/50 | Always Lies | Never Lies | 50/50 | Always Lies |
| Median | 0.44 | 0.21 | 0.51 | 17 | 20 | 23 |
| 97.5 Quantile | 5.63 | 6.81 | 7.42 | 222 | 264 | 311 |
| 99th Quantile | 13.0 | 14.6 | 16.0 | 479 | 575 | 670 |

**Figure 8:**
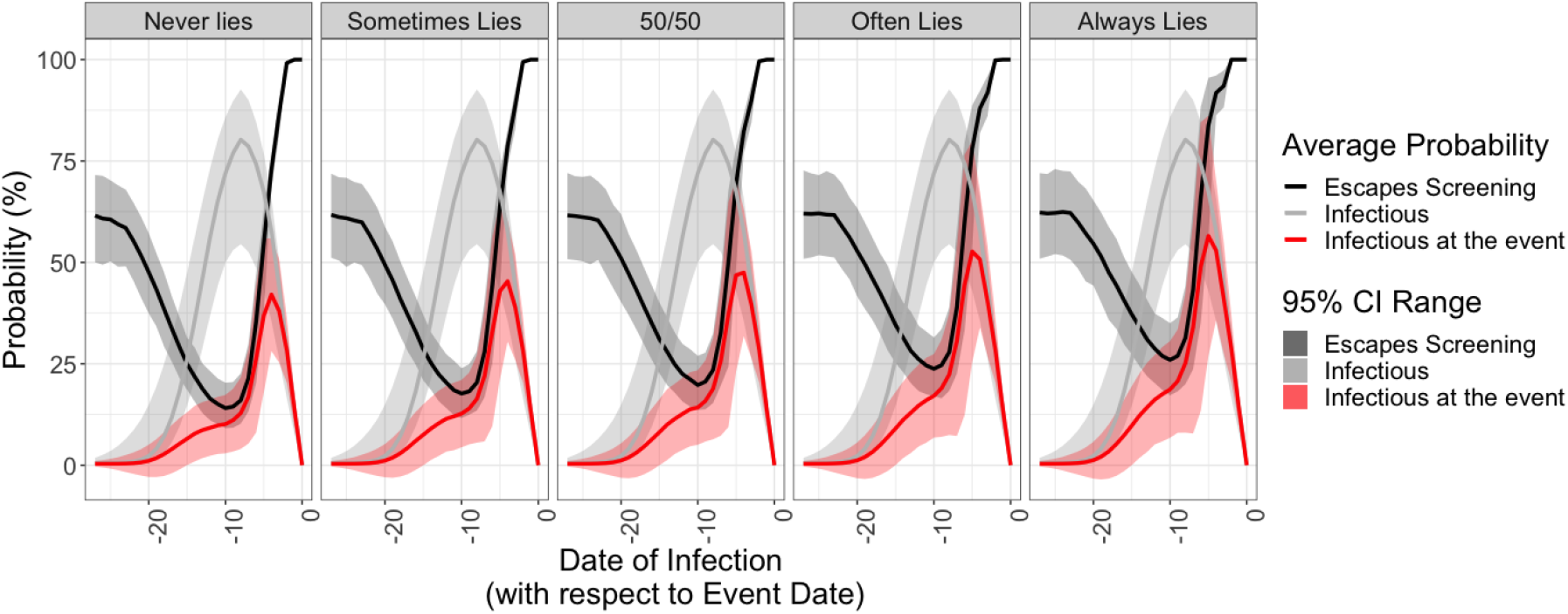
Analysis of the sensitivity of our infectiousness estimation to various values of the probability of lying.

**Figure 9:**
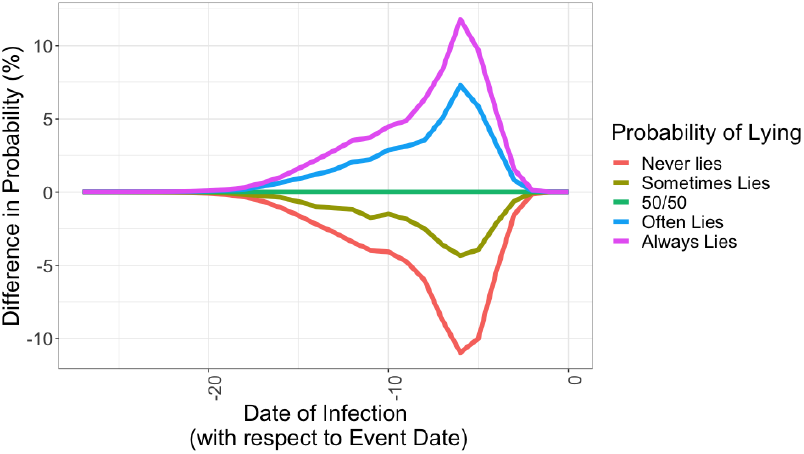
Analysis of the sensitivity of our infectiousness estimation to various values of the probability of lying.

### Mask Efficiency

Another parameter of great importance in the model consists of the effect of the mask efficiency. To this end, we contrast again a scenario with 100% of mandatory mask wearing, but with exhaled efficiency varying from 30, 50, 70, and 90% (the inhaled breathing being fixed efficiency to 50% − the same phenomenon would hold if varying the inhaled breathing efficiency). The results are presented in Table 5. We note the importance sensitivity of the results in the tails of the distribution: the absolute difference (in terms of number of cases) diverges significantly when looking at the 97.5th quantile of the distribution for instance. This highlights the efficiency of masks in limiting superspreading phenomena. We hope to use in particular the results of the CAPACITY study (and behavioural factors such as abidance to mask wearing, preferred type of mask, etc) as well as the growing literature of mask efficiency to be able to refine our estimate of the mask efficiency. In the meantime, in this paper, we use the conservative 70% efficiency (lower bound provided in [67]).

**Table 5:** Sensitivity of the results to Mask efficiency. Note the significant absolute difference in the tails of the distribution.

|  | August 3rd, 2020 |  |  |  | January 18th, 2021 |  |  |  |
| --- | --- | --- | --- | --- | --- | --- | --- | --- |
| Mask Efficiency (%) | 30 | 50 | 70 | 90 | 30 | 50 | 70 | 90 |
| Median | 1.2 | 0.90 | 0.52 | 0.17 | 46.0 | 32.7 | 19.8 | 6.6 |
| 97.5 Quantile | 16.3 | 11.0 | 6.1 | 2.3 | 607 | 440 | 264 | 89 |
| 99th Quantile | 34.6 | 24.0 | 11.6 | 4.9 | 1351 | 959 | 566 | 202 |

### Discussion

In the modelling, we neglect any correlation between ticket holders, however this is unlikely to hold in real life as some might come from the same household. Given participants from the same household would all be rejected if one of them were to test positive, this simplification is likely to be a conservative estimate. Due to the nature of the problem and gaps in what is known about transmission risks, our model does not contend to make precise estimates of the form “ we expect x numbers of cases” as a result of the event. Rather, it should be taken as a means of providing a scale of the risk and is best used to make comparisons, for example against a null model in which the event does not occur but individuals still get infected in the community. As such, our methodology provides a relative quantification of that risk such as “holding the event is expected to increase the number of cases by x% in this pool of participants.” The use of Monte Carlo simulations allow us to account for some uncertainty in our model, and to produce more meaningful risk estimates. As our second goal is to quantify the efficiency of the screening protocol, this pipeline can be run using different screening protocols or testing strategies to determine their efficiency.We also hope to be able to further develop this pipeline through (a) leveraging in-situ data collection and (b) refinement of the models themselves. In particular, using a fully anonymous, post-event questionnaire, we would evaluate participants’ compliance with the screening protocol, as well as with the safety measures at the time of the event. As more events are held, we can quantify the success of our pipeline to provide coverage of the observed number of infections. From the model perspective, we hope to explore further extensions to the current aerosol transmission model, which is based on the Wells-Riley model and so assumes uniform mixing of the quanta in the venue. Using information on the potential induced cases (e.g, their relative seating distance), we aim to refine this model by studying ways of accounting for the spatial heterogeneity of the quanta distribution.

## D. Appendix: Risk Communication

### Vaccine passports and widespread antigen tests − a false sense of security?

The use of vaccine passports for international air travel has ignited significant debate in the UK and even more controversial is their use for entry to mass, live events [12]. Notwithstanding the challenges surrounding operational verification of vaccine certification, the ethical implications of excluding those unable or unwilling to be vaccinated from participating in normal social encounters and the resulting implications for social inequities [13], the use of vaccine certification to permit entry to an event will likely significantly overestimate its safety. Vaccinated individuals may still be infected with SARS-CoV-2. Even antigen-test based screenings of ticket holders prior to an event will likely overestimate the safety of the event as some tests will be falsely negative. The definition of what constitutes an admissible level of risk thus poses a difficult conundrum to the live event industry.

### CAPACITY-UK

The motivating application behind this paper is the CAPACITY study [32]— a partnership between CERTIFIC (a private, remote testing, health status and identify certification service), LiveNation (a live events production company) and Imperial College London − to predict and measure the outcomes of full capacity live events whilst ensuring rigorous abidance by public health and safety measures. Central to this study is the efficiency of pre-event screening by testing all ticket holders using professionally-witnessed rapid at-home antigen tests, and post-event monitoring based on antigen tests, surveys and safety recommendations. Mass rollout of home-based Lateral Flow Testing to all adults in the UK [88] for twice weekly testing ensures that all households will already have the tests available to them. CAPACITY-UK proposes simply for the tests to be professionally witnessed via the CERTIFIC application to overcome the trust issue, verifying that tests have been collected and conducted to the appropriate standard. In addition to testing (which is susceptible to false positives and negatives), the CAPACITY protocol gathers anonymized information on participant vaccination status, regional address, and a few basic questions regarding individual characteristics (see Fig. 10). The purpose of this additional information is to allow the design of a tailored risk estimation model — both at the participant and at the community level. Such risk estimates are central to the protocol: not only are they necessary in the context of informed consent and communicating to the ticket holders their own level of risk so that they may choose to attend the event, they are essential in informing event managers and policy makers on the likelihood of an outbreak. This system potentially allows for the management of full capacity, live events − a crucial parameter for commercial viability of the industry. Moreover, contrary to the issues surrounding vaccination passports, vaccination status would be requested, but not required for attendance − particularly if overall risk of transmission at the event remains within acceptable bounds.

**Figure 10:**
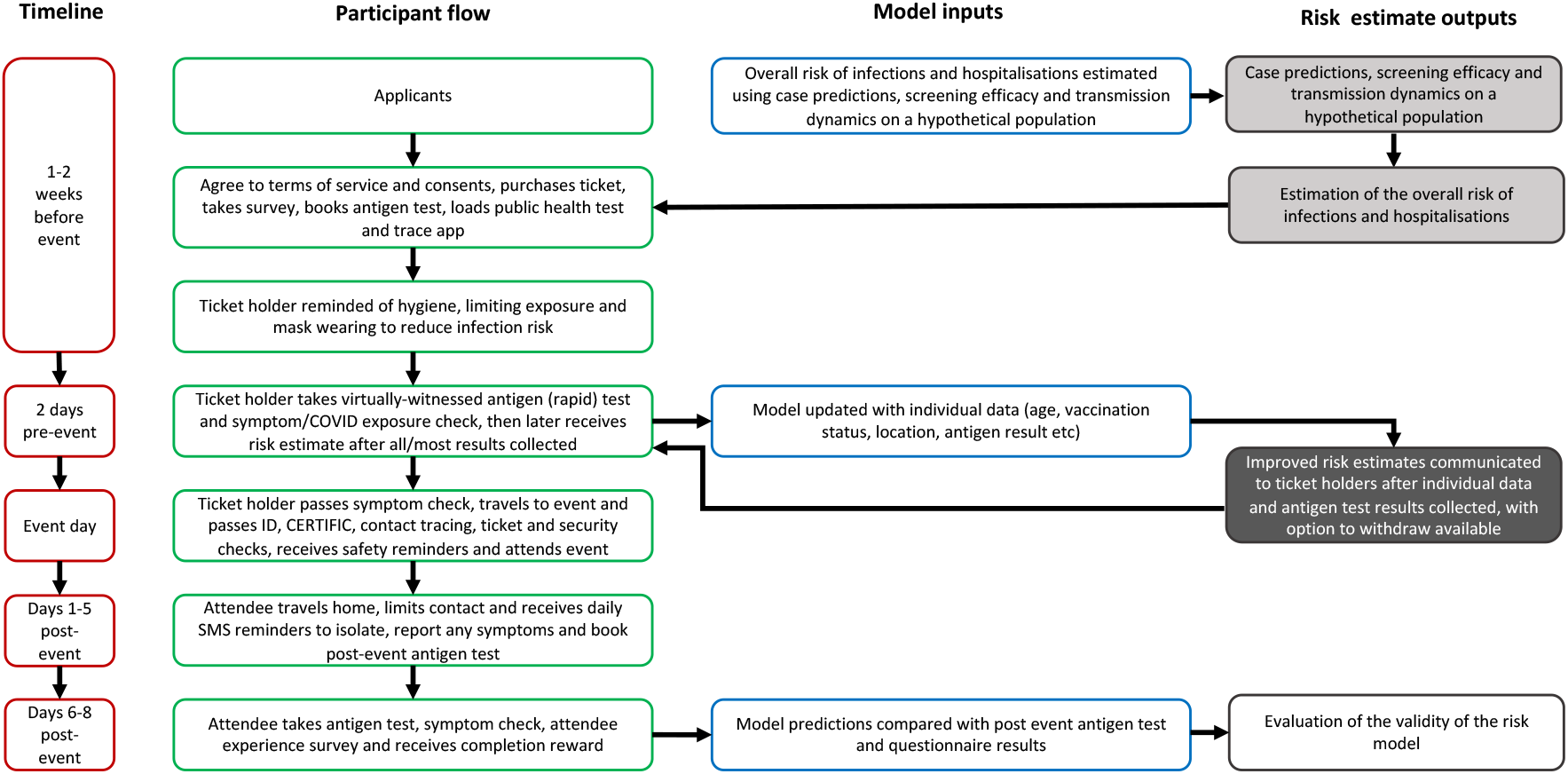
CAPACITY process flow from the ticket holder perspective. On the left hand side in red lined boxes are the timings of various stages in CAPACITY process flow. In the middle the process flow is described from the participant perspective. On the right hand side the interaction between user-supplied data and model-generated risk estimates is described. The certainty in the model output is conveyed through the varying shades of grey (the darker the colour, the more certain the model).

This system potentially allows for the management of full capacity, live events − a crucial parameter for commercial viability of the industry. Moreover, contrary to the issues surrounding vaccination passports, vaccination status would be requested, but not required for attendance − particularly if overall risk of transmission at the event remains within acceptable bounds.

### Risk Communication

The risk estimate is then provided to the participants using a variety of different formats, for better interpretability and communicability of the risk to the general public. Fig 11 shows a few examples of the displays used by the CAPACITY study.

**Figure 11:**
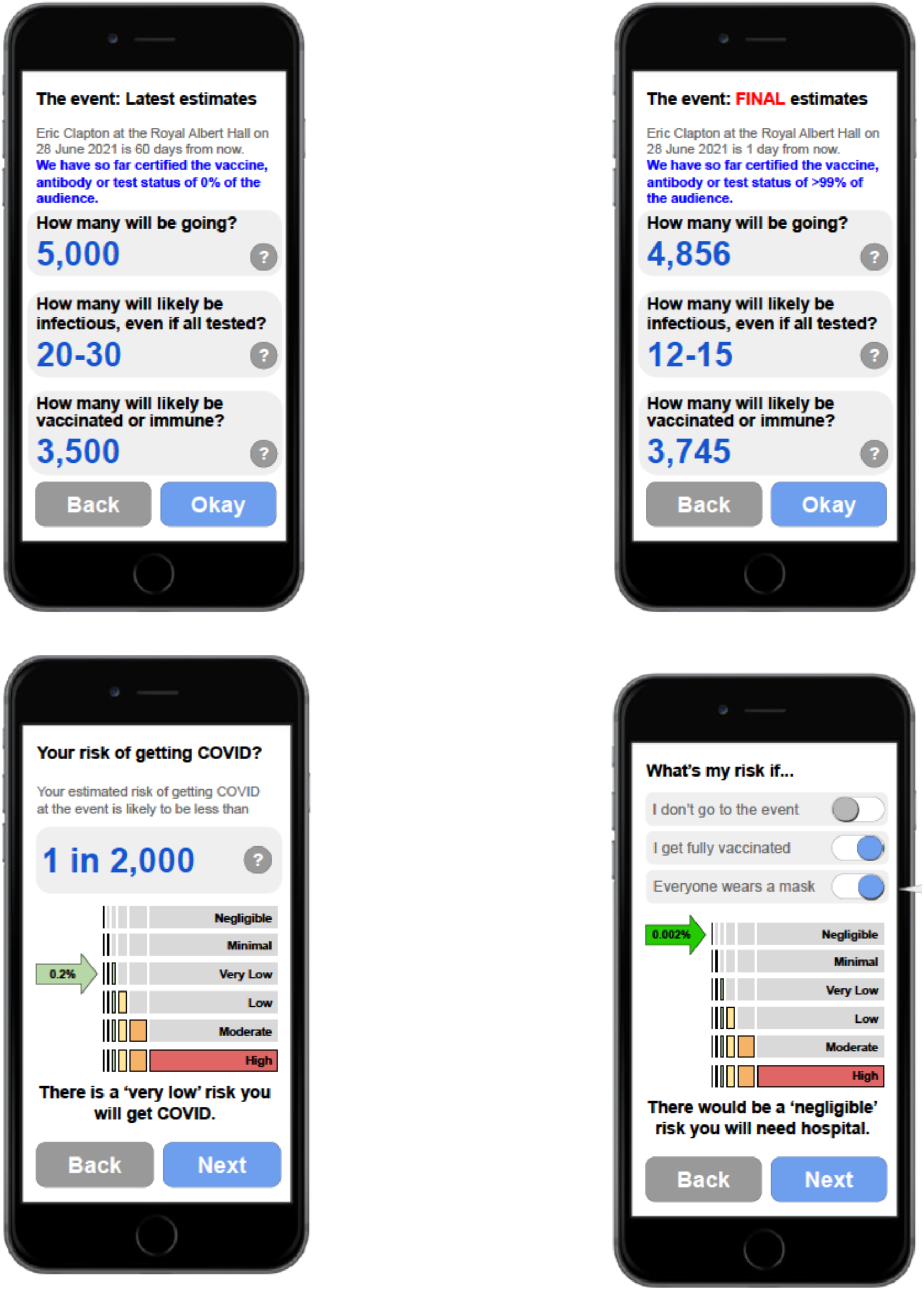
Top left − baseline estimates; Top right − final estimates; Bottom left − individual risk communication; Bottom right − tailored risk scores under different scenarios

## E. Appendix: Reference Tables

**Figure 12:**
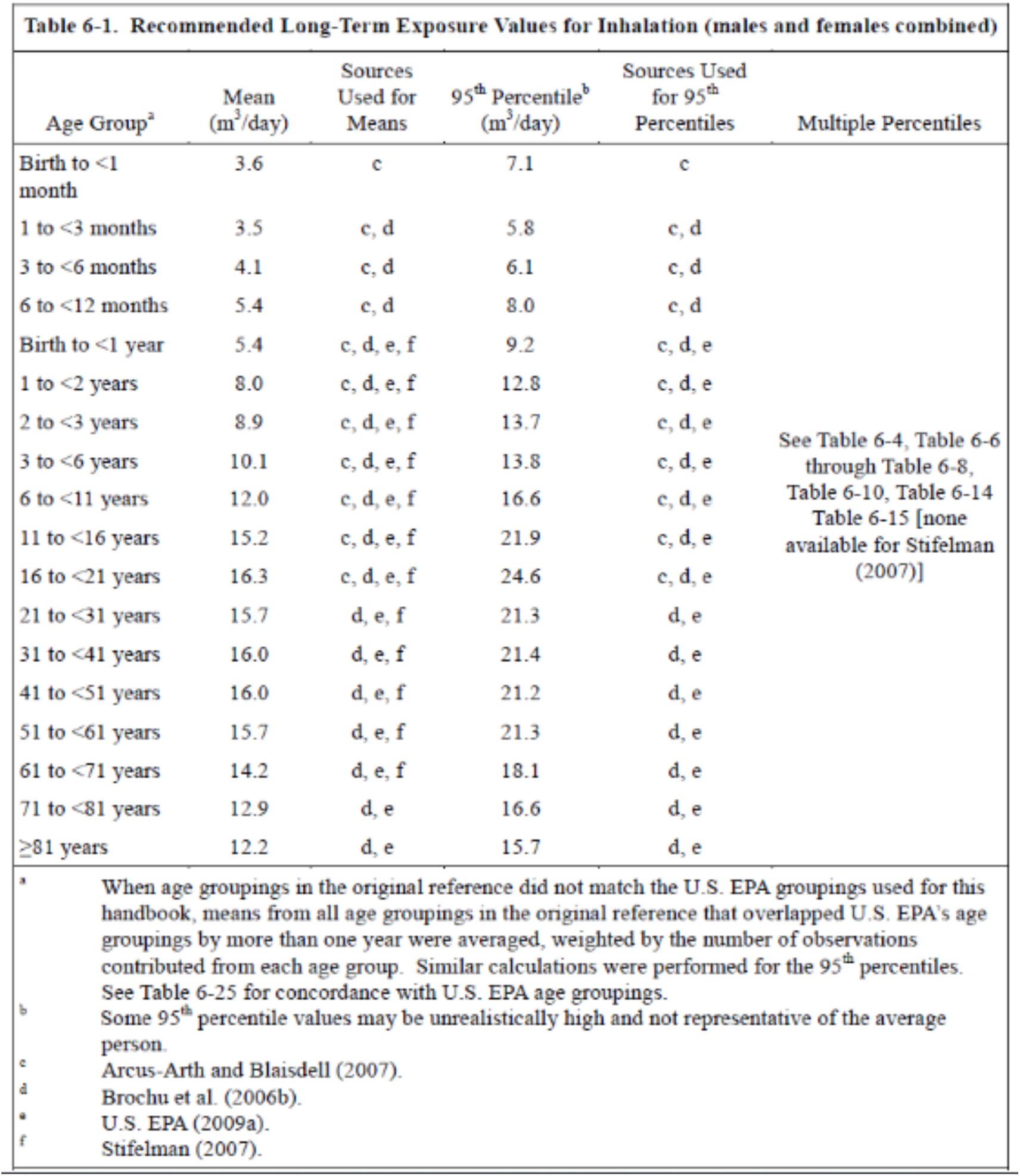
Inhalation Rates. Link to the EPA website:https://www.epa.gov/expobox/exposure-factors-handbook-chapter-6

**Figure 13:**
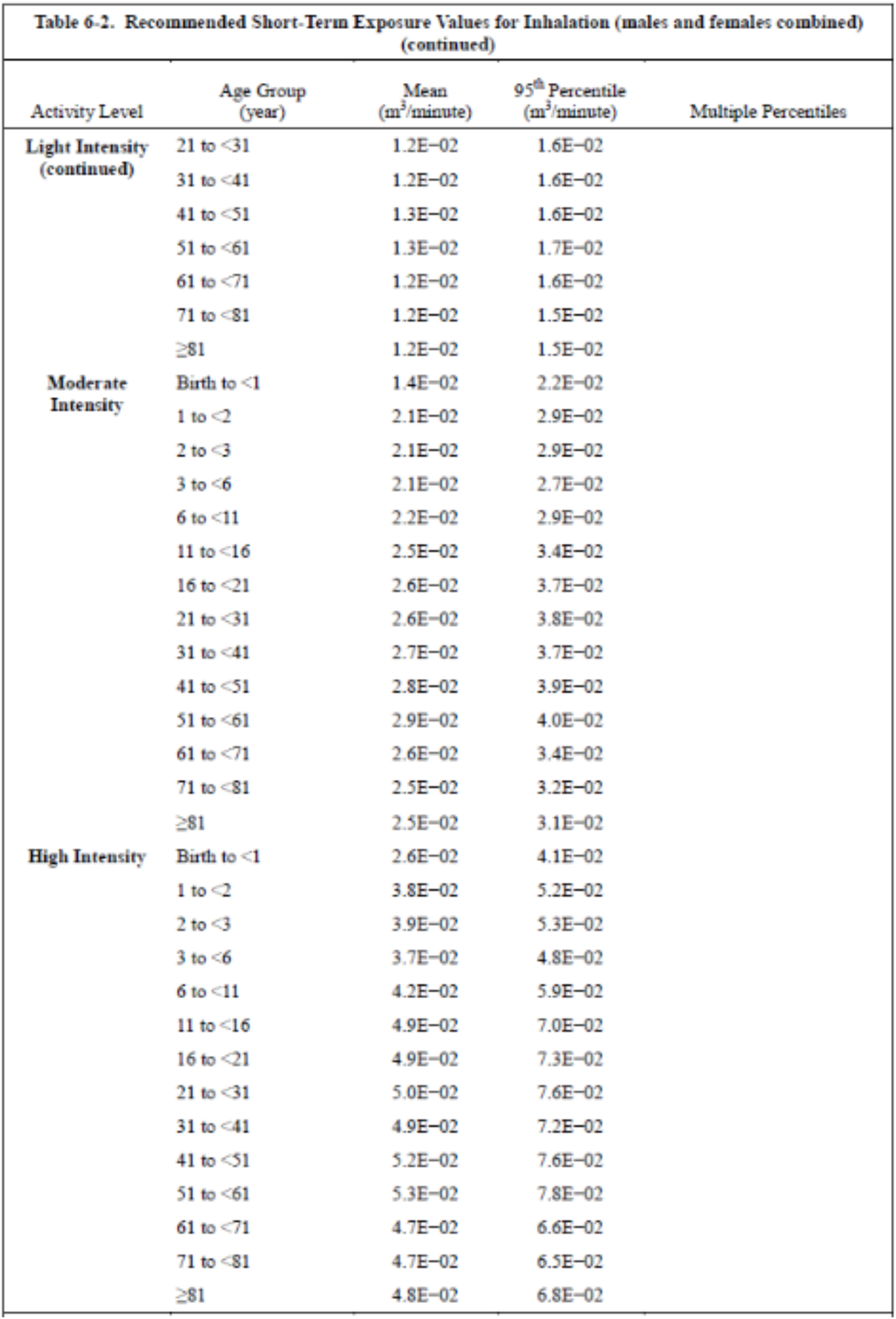
Inhalation Rates. Link to the EPA website:https://www.epa.gov/expobox/exposure-factors-handbook-chapter-6

**Figure 14:**
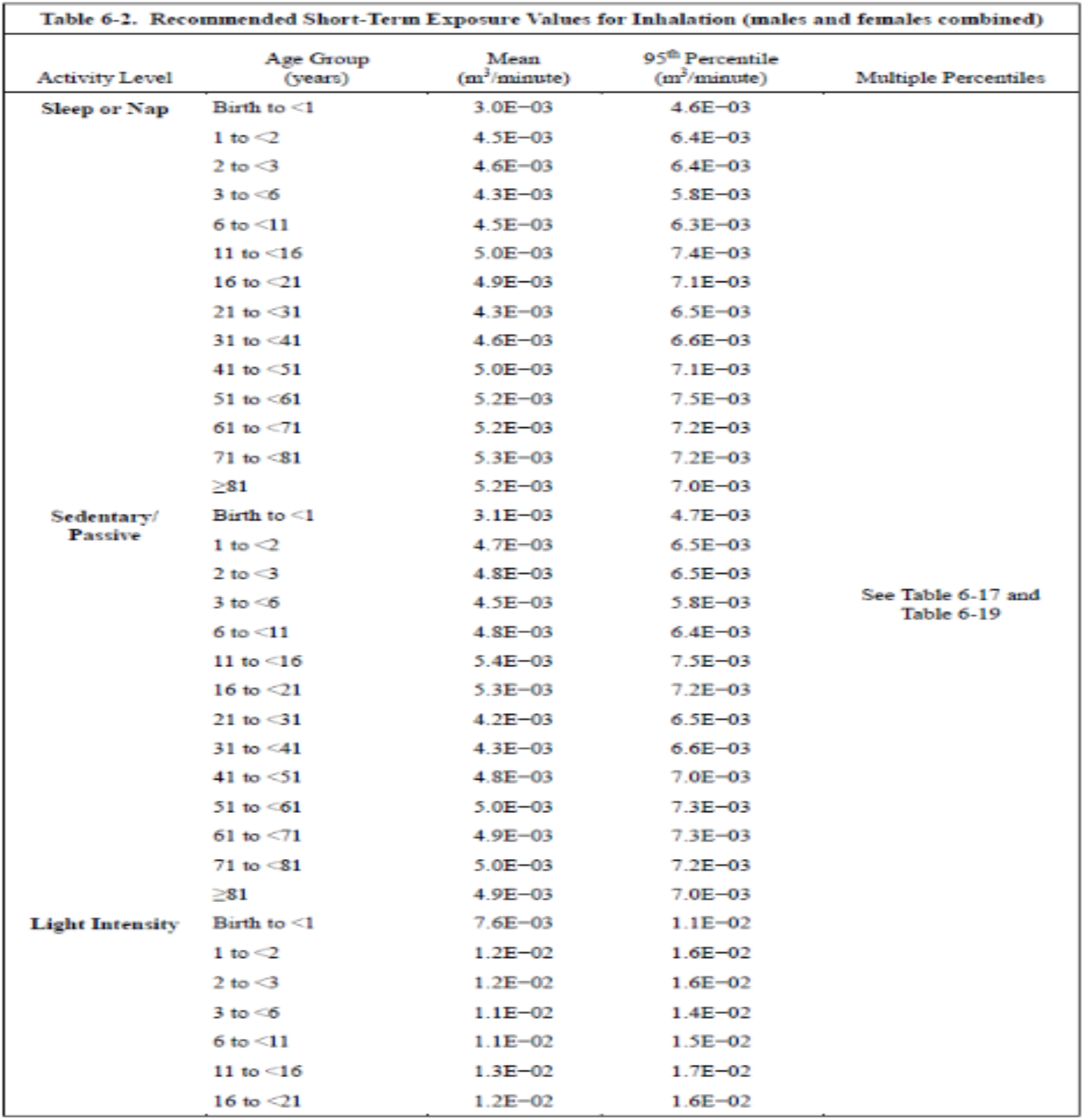
Inhalation Rates. Link to the EPA website:https://www.epa.gov/expobox/exposure-factors-handbook-chapter-6

**Figure 15:**
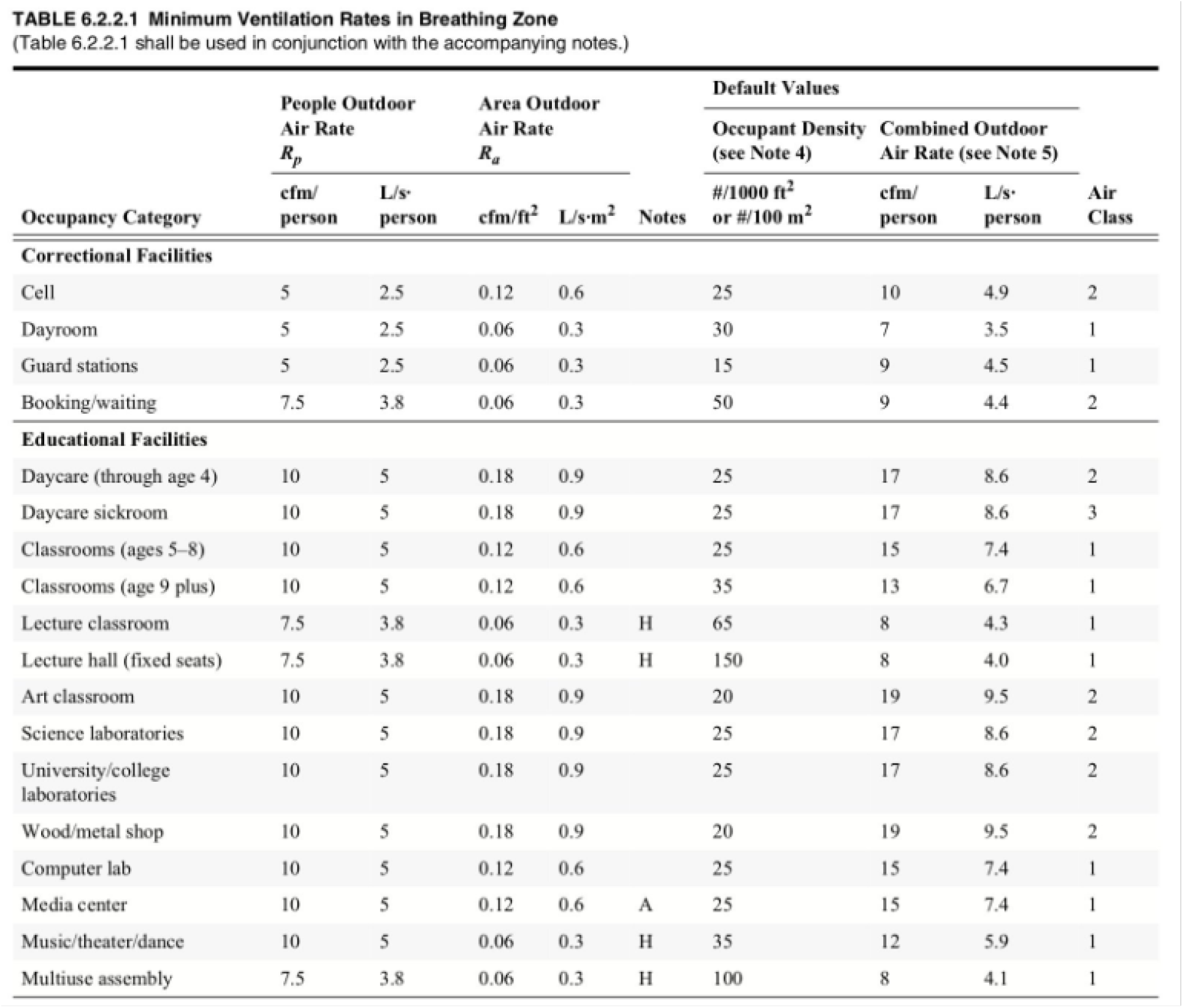
Ventilation Rates taken from the ASHRAE standards ^8^

## Footnotes

1 Source: [Office for National Statistics Survey data](https://www.ons.gov.uk/peoplepopulationandcommunity/healthandsocialcare/conditionsanddiseases/datasets/coronaviruscovid19infectionsurveydata)

2 Link to the prototype: https://homecovidtests.shinyapps.io/aerosol_transmission_model/

3 Data available at the following link: https://github.com/mbevand/covid19-age-stratified-ifr

4 The IFR have been taken from the following data source: https://github.com/mbevand/covid19-age-stratified-ifr

5 This calculator can be found at the following link: https://www.dhs.gov/science-and-technology/sars-airborne-calculator

6 Link to the EPA website:https://www.epa.gov/expobox/exposure-factors-handbook-chapter-6

7 Link to the ASHRAE tables: https://ashrae.iwrapper.com/ASHRAE_PREVIEW_ONLY_STANDARDS/STD_62.1_2019

## Notes

### Competing Interest Statement

JK is a co-founder and Medical Director at Certific.

